# Effectiveness, acceptability and potential harms of peer support for self-harm in non-clinical settings: A systematic review

**DOI:** 10.1101/2021.07.23.21261023

**Authors:** Nada Abou Seif, Rayanne John-Baptiste Bastien, Belinda Wang, Jessica Davies, Mette Isaken, Ellie Ball, Alexandra Pitman, Sarah Rowe

## Abstract

**Background:** Many people who have self-harmed prefer informal sources of support, or support from those with lived experience. However, little is known about whether peer support improves outcomes for people who have self-harmed, and what might be the risks of peer support interventions in non-clinical settings.

**Objectives:** The aims of this review are to examine the effectiveness, acceptability and potential risks of peer support for self-harm, and how these risks might be mitigated.

**Methods:** We searched two bibliographic databases and grey literature for papers published since 2000. We included peer support for self-harm that occurred in voluntary sector organisations, providing one-to-one or group support, or via moderated online peer support forums. We excluded peer support within clinical settings, peer support provided by relatives or friends who self-harm, or peer support from unmoderated online forums. Quality appraisal was conducted on included papers, and study findings were summarised using a narrative synthesis.

**Results:** Ten papers met the inclusion criteria for this review, and most (n=8) were conducted in the United Kingdom. Eight of the papers focused on peer support that was delivered through online mediums and two examined face-to-face self-harm recovery/support groups. Limited conclusions about the effectiveness of peer support interventions for self-harm can be made, as we found no studies comparing this to other treatments or a control group. Peer support for self-harm was found to be acceptable and was viewed as having a range of benefits including a reduction of loneliness, a sense of community and empowerment, improvements in interpersonal skills, and access to information and support. The most commonly perceived risk associated with peer support for self-harm was the potential for triggering self-harm. Other potential risks identified were being re-traumatised by listening to other people’s stories, not having the knowledge or skills to help others, and misunderstandings or disagreements with other peers/group members.

**Conclusions:** Our findings highlighted a range of benefits of being part of a group with very specific shared experiences. This justifies investment in the provision of such resources, supported by safeguards to mitigate the potential risks from peer support interventions. Suggestions include organisations using professional facilitators for groups, including trigger warnings for online forums, and providing regular supervision and training so that peers are prepared and feel confident to support vulnerable people whilst maintaining their own emotional health.

## Introduction

Self-harm is an act in which an individual initiates behaviour (such as self-cutting or ingesting a toxic substance or object), with the intention of causing harm to themselves with a non-fatal outcome [1]. Definitions of self-harm vary according to the degree of suicidal intent, and it is important to note that not all people who practise self-harm feel suicidal when they self-harm [2]. Self-harm is common, particularly in young people, and its prevalence is increasing in many countries around the world [3–5]. Among British 17-year-olds, an estimated 20% of males and 28% of females report self-harm, with white and sexual minority adolescents identified as at particular risk [6]. Self-harm is associated with distress and is the strongest risk factor for suicide [7]. Many young people who self-harm prefer informal sources of support [8], or support from those with lived experience available through voluntary sector organisations and online forums.

Peer support interventions are becoming increasingly adopted worldwide within mental health services and third sector organisations (voluntary organisations, community organisations and charities) [9]. Although studies of peer support for mental health problems suggest it is associated with positive effects on hope, recovery and quality of life, the effects on other outcomes such as symptoms, hospitalisation and satisfaction are inconclusive [10]. Peer support may have different benefits and harms depending on the population and setting in which it is used. Peer-reviewed research on peer support for people that self-harm is lacking, and little is known about the provision, quality, effectiveness and acceptability of these resources for young people or adults, or the potential harms. This hampers the planning of appropriate service provision for people who have self-harmed.

In this systematic review of the quantitative and qualitative literature, we aimed to address the following research questions:

1. What evidence exists for the availability and effectiveness of peer support for self-harm?
2. What evidence exists on the risks of peer support for self-harm where risk, safety or normalisation may present concerns?
3. What evidence exists on how the risks identified through the review could be mitigated?

## Methods

Our systematic review followed PRISMA guidelines and was registered on the PROSPERO international prospective register of systematic reviews (CRD42021235441).

For the purpose of this review and in consultation with Samaritans, we used the term ‘self-harm’ to describe self-harm behaviour where there was no suicidal intent, ‘suicide attempts’ where there was suicidal intent, and ‘self-harm/suicide attempts’ where both applied or where this was unspecified or unclear. This was because we wanted to define the client group of interest very tightly, recognising that the provision, effectiveness, acceptability and potential harms of peer support for suicidal self-harm is likely to differ from that provided for non-suicidal self-harm. We also defined ‘peer support’ for self-harm as any support provided in non-clinical settings by individuals with lived experience of self-harm. We excluded peer support within clinical settings, as recommendations from a recent parliamentary inquiry were to invest in community-based preventative services, including low level preventative support based on peer support models [11]. We also excluded peer support provided by relatives or friends who self-harm, or peer support from people posting content about self-harm on the internet solely in a personal capacity as these models would not benefit from funding.

Our definition therefore included: peer support provided by individuals working for voluntary sector organisations (but not formal healthcare services) providing one-to-one or group support, or via moderated online peer support forums.

## Searches

Searches were conducted on MEDLINE® (Ovid) <1946 to February 15, 2021> and PsycINFO (Ovid) <1806 to February Week 2 2021>. Search terms were developed with the input of a lived experience researcher (JD) and in collaboration with the Samaritans team (EB and MI). Search terms covered keywords relevant to self-harm and both online and face-to-face peer support. These were combined into a single search string using the appropriate Boolean operators (Supplementary material 1).

We also conducted searches for grey literature on both Open Grey and Google using the approach suggested in recent scoping reviews (Pham et al 2014 https://www.ncbi.nlm.nih.gov/pmc/articles/PMC4491356/). For the Google searches, we made an *a priori* decision to screen only the first 100 results to reflect a balance of relevance and time taken to screen each hit [12].

We searched websites for UK charity organisations such as Mind and Harmless and contacted each organisation to request any relevant publications on peer support. The list of organisations was comprised from suggestions by Samaritans and our team’s lived experience researcher (Supplementary material 2).

Studies were included if they:

1. described the provision, quality, effectiveness, and acceptability of peer support for self-harm
2. related specifically to self-harm, regardless of suicidal intent
3. were published in English
4. were published from the year 2000 onwards
5. used quantitative, qualitative or mixed methods

We did not set any restrictions on age group, population, study design, or whether publications were peer reviewed. We also included studies with or without a comparator group, as we felt this was particularly important in capturing acceptability of the peer support intervention.

We excluded studies that:

1. focused on suicide prevention without investigating self-harm specifically
2. described peer support in clinical settings
3. described peer support provided by relatives or friends
4. described peer support from people posting content on the internet solely in a personal capacity
5. peer support taking place on unmoderated online mediums (eg, unmoderated forums)

## Data extraction

### Screening and selection of studies

We used Covidence systematic review online software (www.covidence.org) to import references from our search engines for screening titles and abstracts, and deduplicate. Two reviewers screened all titles and abstracts independently. Four reviewers screened the full-text articles independently to determine their suitability for inclusion and a randomly selected 10% of these were second screened by an independent reviewer. Reference lists of included papers and relevant systematic reviews were checked for relevant papers. Any disagreement between the reviewers over the eligibility of studies was reviewed by a third and fourth reviewer (SR and AP) and resolved through discussion. We calculated inter-rater agreement at each stage of the review screening process to assess the consistency of raters’ decisions. We used the accepted value of 0.8 as the threshold for good inter-rater agreement [13], resolving screening disagreements where values fell below 0.8 resolved through discussions with a third and fourth reviewer.

### Data extraction

Information on the following variables were extracted from all of the papers: study titles, authors, study type, country of origin, year of publication, population, demographics (including age, sex, ethnicity), type of self-harm (suicidal, non-suicidal, both or not specified) details of peer support intervention (nature, description, duration, source of provision), outcome measures, change scores or themes relevant to peer support intervention, risks or harmful effects, and mitigation of risks or harmful effects. A second reviewer independently checked data extraction.

The outcome measures in relation to each of our review questions were:

1. Mean reduction in self-harm behaviours post peer support intervention
2. Changes in mean questionnaire scores, or themes relevant to risk/harmful effects of self-harm peer support interventions as derived from qualitative research
3. Ways in which the risks identified in research question 2 might be mitigated, as established using the results and discussions sections of all included papers

### Quality appraisal

One reviewer independently assessed the quality of each included paper, and a randomly selected sub-sample of 10% of included papers was independently assessed for study quality by a second researcher.

For quantitative peer-reviewed published papers we used the Grading of Recommendations, Assessment, Development, and Evaluation (GRADE), which rates papers on five domains: risk of bias, imprecision, inconsistency, indirectness, and publication bias. A certainty/quality rating is assigned to the evidence ranging from very low (the true effect is likely to be substantially different from the estimated effect) to high (we are confident that the effect of the study reflects the actual effect) [14].

For qualitative studies we used the Critical Appraisals Skill Programme (CASP) Qualitative Checklist, which examines whether there is sufficient description and justification of the chosen methods of data collection, sampling, and analytical approach, as well as whether sufficient attention was given to ethics and the role of the researchers involved [15].

For studies using a mixed method design we used the Mixed Methods Appraisal Tool (MMAT) version 2018 [16], which includes a checklist to appraise the methodological quality for qualitative, quantitative and specifically mixed methods studies. MMAT examines whether the rationale of using a mixed methods design is appropriate and whether the different components of the study are incorporated constructively to answer the research question.

For non-academic papers (including grey literature), we used the NESTA’s Standard of Evidence model to assess the quality of each source of information [17]. This considers criteria such as “Have others proved the same?” and “Can this be replicated elsewhere?” to judge whether the innovation described has evidence of benefits or harms.

### Data synthesis

Anticipating a heterogeneous range of papers, we used a narrative synthesis to summarise themes relevant to our review questions. In team discussions, including Samaritans team members and our lived experience researcher, we explored reflexivity in our interpretation of the findings to ensure that our inferences regarding recommendations for practice were appropriate, acceptable and relevant.

## Results

Our MEDLINE and PsycINFO searches identified 31,667 records, with an additional 35 records identified through OpenGrey and Google searches (Figure 1). After the removal of duplicates, a total of 26,523 titles and abstracts were screened. Of those, 28 full-text articles were assessed for eligibility, from which a total of nine studies were identified as eligible for inclusion in our final synthesis. A further 35 records were identified from non-profit organisations, which we reduced to seven following deduplication and title and abstract screening. After full-text screening, one of these records was judged to meet eligibility criteria for inclusion in our final analysis based on its specific focus on online peer support for young people self-harming [18]. Other records from non-profit organisations were excluded due to peer support being offered for difficulties not limited to self-harm and not fitting our description of a peer support intervention. Inter-rater agreement was high for screening titles and abstracts (99%) but went down to 75% for the full-text screening. One paper required discussion with a third reviewer before a consensus could be reached on its inclusion in the review and inter-rater agreement over 0.8 could be achieved.

**Figure 1.**
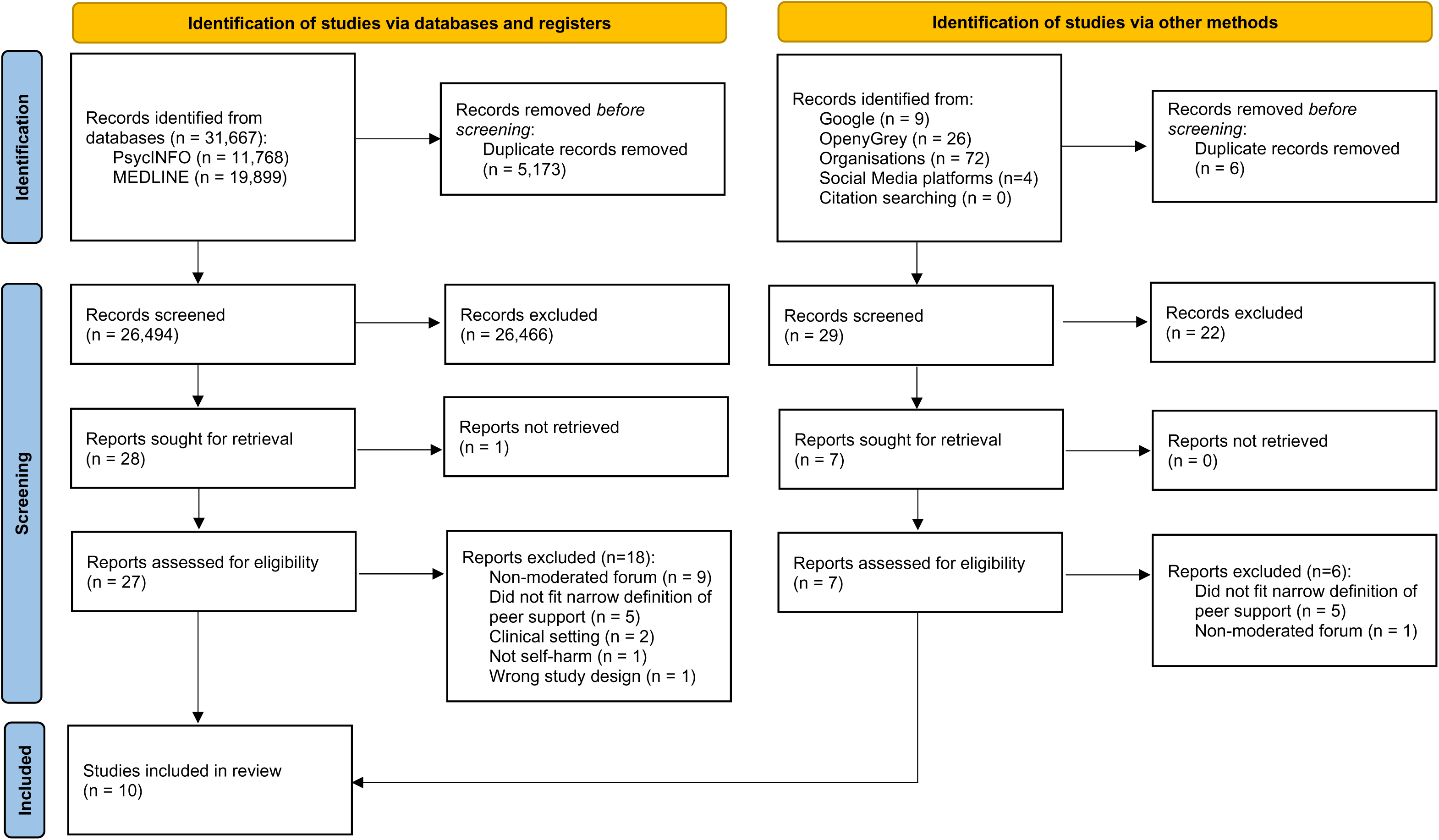
PRISMA flow diagram

### Description of studies

Of the ten studies included in our narrative synthesis, eight were conducted in the UK, one in the US, and one paper combined data from a range of mental health organisations in the UK, Italy, Slovenia, and Denmark [18]. Of the included studies, eight were of qualitative design and two used mixed methods (Tables 1A and 1B). Notably, no randomised controlled trials or other trials investigating effectiveness met our inclusion criteria. In eight of the studies, peer support was delivered through online mediums, such as self-harm forums or message boards (n=5), online recovery groups (n=1), social media (n=1), or a variety of platforms, including group chats, online group discussions and Facebook groups (n=1). The remaining two studies examined face-to-face self-harm recovery/support groups.

As per our inclusion criteria and research focus, all studies focused on individuals who self-harm, with five studies focusing specifically on young people and/or young adults who self-harm [18–22]. Studies included a wide range of sample sizes, ranging from n=7 [23] to n=102 [24] participants. However, the UTRIP (2012) report was an outlier, as they included a range of samples sizes, and it wasn’t clear if they used the same participants for different stages of their surveys (Table 1B). Although most studies did not report ethnicity, where ethnicity was reported (n=3), individuals self-defining as White constituted 100% of the sample in two studies [23, 25], and 70% of one other [19]. Females made up at least 80% of the samples across all studies. The majority of studies investigating online peer support sampled individuals within a young age range (16 to 25 years), apart from Haberstroh & Moyer (2012), in which the sample had a mean age of 36 years. The two studies investigating face-to-face peer support had a sample with a mean age of 36 and 46 years, respectively [23, 26].

### Quality of included studies

All eight qualitative studies included were judged to be of high quality, scoring 8 or above out of 10 on the CASP (Supplementary material 3A). However, studies tended to be unclear on how they addressed the relationship between the researchers and participants. Nevertheless, the other domains of CASP were judged to be addressed adequately in most studies. Of the two mixed methods studies, one [27] was scored as 11 out of 15 on the MMAT (as used for mixed methods research), with the domains for qualitative design judged as well addressed, but less so for the quantitative design (Supplementary material 3B). The other mixed methods study [18] was appraised using NESTA (as used for grey literature) and only rated as level 2, as the lack of comparator groups meant that the effects of the intervention could not be separated from other influences (Supplementary material 3C).

### Findings

The ten included studies identified a range of views on the acceptability and perceived value of peer support for self-harm, both from the perspective of those using the service and those providing it, along with descriptions of the mode of provision (Table 1A and 1B). We did not identify any studies reporting the effectiveness of peer support for self-harm, nor studies presenting an overview of the provision of peer support for self-harm nationally or internationally.

## Q1. Mode of peer support, acceptability, and effectiveness

### Face-to-face peer support

Both the studies evaluating face-to-face peer support focused on self-harm support groups, and both reported on the experiences of members of more than one support group. Participants in the Boyce et al. (2018) study discussed how members’ experiences prior to joining the group had been primarily negative and characterised by isolation, stigma, and shame [26]. Conversely, participants in both studies viewed their self-harm support groups as a safe space, where they felt accepted and understood (Table 2). Their shared experiences with other members of the group made participants feel that the support they were receiving in this setting was more “genuine” than that on offer from professionals or family/friends. Many of the group members in both studies reported a reduction in self-harm following group membership. Participants described other positive changes that they attributed to group membership, such as friendship and decreased isolation, improvements in self-awareness, mood, and interpersonal skills. They also reported that they derived a sense of empowerment and self-worth through witnessing and supporting each other’s struggles and successes. Peer support group leaders reported positive experiences in relation to their sense of autonomy in running the group [23]. These findings suggest that self-harm support groups are perceived by members as valuable peer support in helping to manage self-harm and are also acceptable to its members.

**Table 2:**
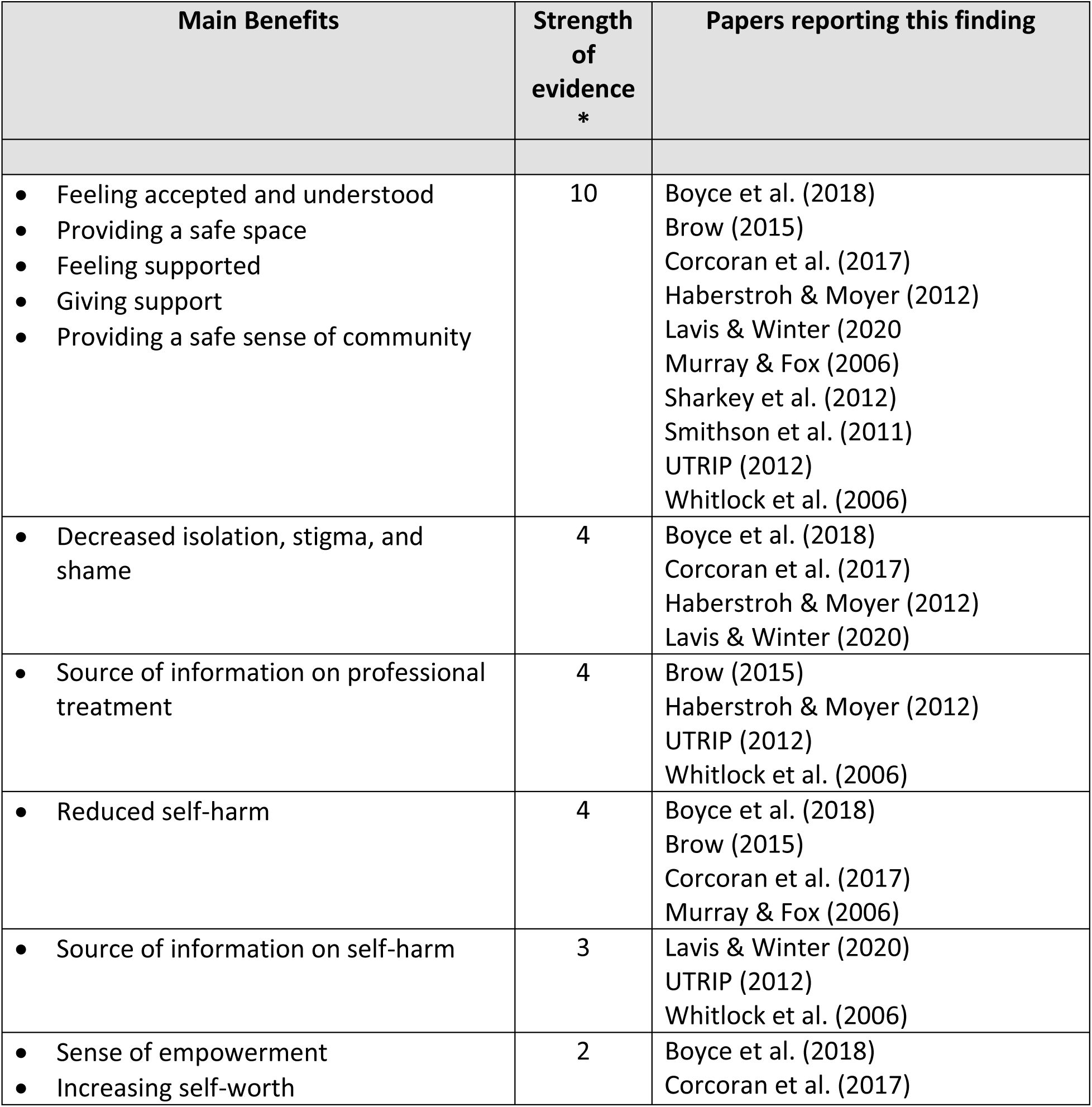
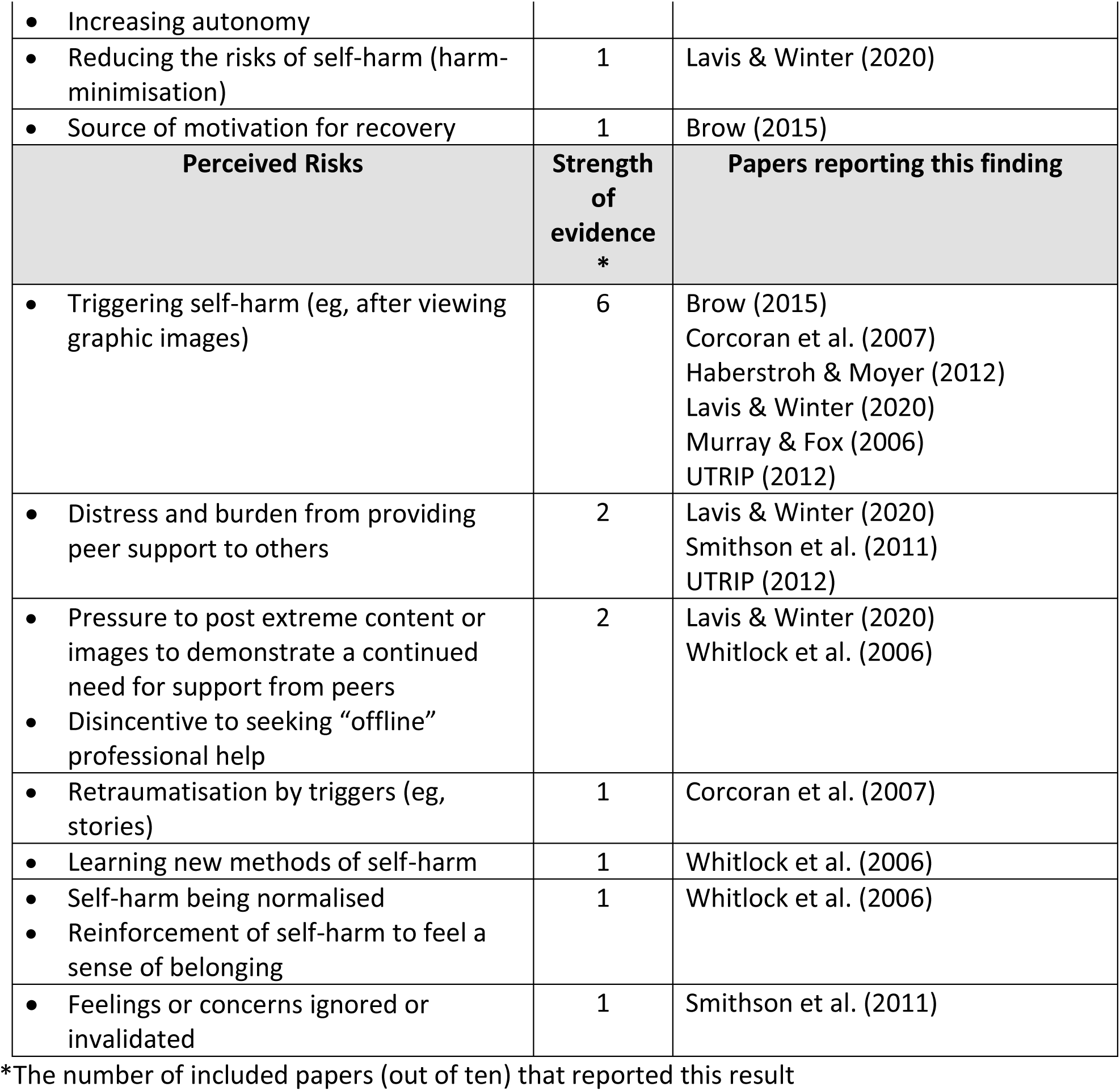
Summary of the main benefits and risks identified in included studies

### Online peer support

Most studies of online moderated peer support investigated self-injury message boards or forums (n=5/8; [19, 21, 24, 27, 28]. Of the remaining three studies, one explored an online recovery group, which consisted of individuals referred to the group by self-injury professionals [25], one investigated young people’s engagement with self-harm content on social media, such as Reddit, Instagram and Twitter [20], and one investigated online peer support services for young people who have self-harmed provided by a range of organisations across Denmark, Italy, Slovenia, and the UK [18].

Similar to the findings for face-to-face peer support, users of online peer support reported that their previous experiences of conventional (ie, professional or non-peer) support were characterised by poor treatment and stigma, which drove them to seek alternative options [25] [20]. The key advantages perceived by participants in these online mediums included providing and receiving support from those with similar lived experiences [20, 24, 25, 27]. Advantages were also perceived in gaining access to useful information on self-harm [18, 20, 27], such as how to self-harm safely [20], how to conceal self-harm (ie, methods of concealing cuts and scars, how to seek treatment [18, 27], and less anxiety around people finding out about their self-harming behaviour [27]. In one study, the online group was also viewed as a useful supplement to counselling [25]. However, a small proportion of participants (n=12/74; 16%) in the study published by Murray & Fox (2006) reported that they used the forum with the intention of being triggered to self-harm [24]. These participants described feeling competitive about their self-harm when reading posts, with some viewing their self-harm as “inadequate” and some deliberately reading discussions when they felt the urge to self-harm [24].

As no trials of online peer support met our inclusion criteria, effectiveness could not be established. In studies where participants were asked about the perceived effectiveness of online peer support in reducing their self-harm behaviours, 41-50% reported a decrease in self-harm, which they attributed to group membership [19, 24]. In the Murray & Fox (2006) study, 46% reported no change in their self-harm, and 10% reported an increase [24]. The sense of safety and community stemming from online peer support made users feel supported by people who understand their experiences of self-harm, and less alone [18, 19, 24, 25]. Added perceived benefits included the sense that online peer support increased participants’ motivation to recover [19], prompted help-seeking for professional (ie, formal) support [19, 25], and was accessed quickly and easily as a source of information [18].

## Q2. Risks and mitigation

### Face-to-face peer support

Risks discussed in the Corcoran et al. (2007) study included the potential for members to be re-traumatized through listening to each other’s stories, as well as the risk of triggering self-injury through learning new methods within the group [23]. The article included suggestions as to ways in which this could be mitigated, including the use of a professional facilitator who could establish clear and healthy boundaries within the group, as well as supplementing the group intervention with individual support.

### Online peer support

Included articles identified three main risks perceived with online peer support interventions. First, the most frequently documented perceived risk identified from online peer support for self-harm was the risk of triggering self-harm behaviour [18–20, 25]. This could arise through: 1) unmoderated sharing of triggering content, such as graphic images, distressing stories, or new methods of self-harm or self-harm concealment methods [25, 27], 2) having to prove continuously a need for help by posting more extreme content or images in order to sustain online peer support [20], 3) a desire to belong [27], or 4) the normalisation and reinforcement (sometimes seen as an encouragement) of self-injurious behaviour [27]. The second most common potential risk identified was that the use of these mediums could isolate members from the “offline” world and hinder them from seeking professional and/or offline help [20, 27]. The third most common potential risk identified was that online peer support might negatively impact the wellbeing of peer supporters due to the distress and burden associated with hearing others’ stories and attempting to help [18, 20] as well as feeling ignored [21], misunderstood or being involved in a disagreement with other members [18]. Additionally, there was the potential for young people to feel overwhelmed while supporting others, as they may lack the skills or knowledge of how best to help[18].

One way to mitigate these risks is to have an online moderator who enforces boundaries between members and overt “no triggering” rules [18, 21, 25]. However, moderators are sometimes individuals who self-harm or who have self-harmed, and there is a risk of this involvement in being triggering for them too [25]. Where moderators have lived experience of self-harm, it was suggested that they should be individuals who have established recovery and are well-supported outside the online group [18, 25]. Some studies also highlighted the potential for clinicians to play an important adjunctive role in peer support for self-harm in two ways: clinicians may act as moderators where peer support is offered in clinical settings but should also inquire about patients’ engagement with peer support for self-harm offered online [18, 19], so that they can support them in using forums constructively. It was also suggested that online mediums should also signpost to offline resources to provide a safety net should other support be needed [20]. One study also alluded to the potential benefits of smaller group size in online forums, which meant users were less likely to feel ignored [21].

## Discussion

The main findings of this systematic review were that only a few studies have investigated the provision, quality, acceptability, effectiveness or potential harms of peer interventions for self-harm, whether in the published or grey literature. Of the studies available, none have evaluated the effectiveness of peer support for self-harm, which limits conclusions about its impact on key outcomes such as distress, stigma, depressive symptoms, suicidal ideation, suicide attempt, and suicide, as well as objective measurement of the risk of potential adverse effects.

However, we did identify useful literature describing the perceptions of those who use peer support for self-harm in relation to its acceptability and potential harms. These studies described a range of perceived social benefits, including a reduction in loneliness and the gaining of a sense of community and interpersonal skills, and also described a range of emotional benefits such as the sense of being able to help others, an opportunity to vent frustrations, and the provision of access to information and support. Additionally, these studies described perceived clinical benefits, including a reduction in the frequency and severity of self-harm, improvements in mood, and an increase in the practice of safer methods of self-harm. No participants described feeling less suicidal as a consequence of using peer support for self-harm. However, this question would better addressed quantitatively using validated measures of suicidality. Many participants described how stigmatised and ashamed they had been made to feel when using more conventional sources of support for self-harm, contrasting this with their positive experiences of the community they had encountered through peer support.

Some important risks were also identified, including the potential for peer support to cause vicarious trauma or trigger self-harm behaviour in those listening to others’ stories or viewing others’ scars, as well as the potential for psychological processes such as reinforcement of self-harm, imitating others’ self-harming behaviours, comparing extent of injuries, or self-harming in order to fit in with peers. There were also concerns expressed about the burden on those participating in peer support in having to support others despite not having appropriate training. Specific potential risks of online peer support were the difficulties of monitoring whether participants felt safe, and the potential for participants to rely on online support over formal sources of support, where this might otherwise be indicated. However, a number of valuable suggestions were made as to how these identified risks might be mitigated, including providing professional facilitators for groups and trigger warnings, and ensuring that peers who take on moderating roles feel well-supported themselves.

## Strengths and limitations

Our systematic review had a number of key strengths, reinforcing the confidence of practitioners and policymakers when applying our findings to practice. We gained the input of a researcher with lived experience, a mental health professional, a health psychologist and of Samaritans when identifying search terms for this review and used these professional networks to contact key voluntary sector organisations and self-harm groups in order to identify grey literature. We also used Twitter for suggestions on published and unpublished literature on potential harms, in order to balance our study and counter potential publication bias. Our quality assessment of included studies used standardised tools and were judged to be of high quality. Our summary of the potential benefits and harms of peer support for self-harm presented a balanced account of the key considerations described in this published and unpublished literature when implementing peer support for self-harm, including key risks and mitigation recommendations.

In only searching two databases we may have missed studies published in other journals, but our use of MEDLINE and PsycINFO was intended to focus our review on clinical findings. Our focus on non-clinical settings may have limited the lessons to be learned about other benefits of peer support for self-harm, but this specific focus was intended to address a research and policy gap in relation to non-clinical settings. We acknowledge that although our search of the published literature included international studies, our exclusion of non-English language studies will have biased our report to reflect primarily the experiences of high-income countries. We also acknowledge that our search for grey literature reflected primarily UK-based organisations, given the location of the research team and funder, and this report might therefore be of less relevance to non-UK settings. We did not identify any trials, despite our comprehensive search terms, which meant that we could not present evidence of effectiveness. We also noted that the samples in included studies tended to underrepresent the experiences of people from minority ethnic groups and this may reflect sampling biases within those studies. We acknowledge that we may not have contacted the full range of experts in the field, who may have unpublished data not represented in this review and did not contact authors to clarify any queries over the presentation of data.

In this review, our exclusive focus on peer support for self-harm may have neglected a wider perspective in which peer support is compared directly to other forms of support for self-harm in terms of their relative acceptability. The findings of an Australian survey of young people who have self-harmed is particularly striking in this respect [29]. This study aimed to explore the attitudes of young people who have self-harmed towards the use of online help for self-injury, as a means of informing future service delivery. Survey responses from 457 young people who have self-injured identified preferences for future online help-seeking, the rationale for which included gaining information and guidance, reducing isolation, a preference for an online culture, facilitation of help-seeking, easy access to support, and advantages of privacy. Of all sources of online support listed (eg, texting, gaming, direct links to professionals, self-help, and peer support) the most popular option was contact with a professional via instant messaging. Professional help therefore appeared to be preferred to peer support within the online context, highlighting the importance of considering hybrid sources of support.

## Future research

The main gaps in research that we identified were studies describing the effectiveness or cost-effectiveness of peer support for self-harm, as we did not identify any trials or economic analyses. The published and unpublished studies we found suggested that peer support for self-harm is an acceptable approach to a specific sub-set of people who have self-harmed and that they, and the professionals who support them, show a strong awareness of the potential risks and mitigations in order to provide a safe service. Although this review identified the triggering of self-harm as being the most common potential risk attached to peer support, no studies evaluated whether the benefits of peer support interventions for self-harm outweigh the risks. Clinical trials and large-scale observational studies are required to measure both positive and adverse effects so we can build on our subjective understanding of peer support for self-harm. We also need cost-effectiveness analyses that take a wide societal perspective, taking into account the potential for clinical benefits and adverse effects, the costs and benefits to carers, the health service, and emergency services, as well as use of other support and treatment options.

We identified no studies describing the geographical provision of peer support for self-harm, nor an overview of current online provision. This is needed to ascertain whether there are disparities in provision for certain geographical areas or for certain digitally excluded groups. In particular, we hope that future studies explore the acceptability of peer support for people from ethnic minorities who self-harm, and that the lack of trial evidence might be addressed through the conduct and publication of controlled trials. We would recommend that regular updates of this review will inform updated recommendations on the provision, quality, acceptability, effectiveness and potential harms of peer support for self-harm.

## Policy implications

These findings suggest that the provision of peer support for self-harm is acceptable and valued by some people who have self-harmed, but that participants also perceived specific risks and ways to mitigate these. The range of benefits described suggest that there would be value in implementing further peer support services for people who have self-harmed, provided that service planning included a careful consideration of risk management. The generally young age of the samples described in this review, typically aged 16 to 25 years, suggests that this would be welcomed by adolescents who self-harm, given their general preference for peer support over formal support [8]. The stigma and shame experienced when accessing professional support was contrasted with the more accepting experience of using peer support. This has important policy implications, given that stigma has been identified as a key barrier to seeking professional support for mental health problems among young people [30]. Online peer support was viewed positively as a way of promoting help-seeking for professional support and may be a critical stage in the process of recognising a need for support and identifying the most acceptable routes into professional support.

The suggestions made around risk mitigation processes suggest a key role for training and supporting group facilitators and online group moderators, both in relation to the processes they follow when providing peer support (policies on triggering content, ground rules about expectations in showing respect in interactions) but also in relation to supporting their emotional needs. The provision of regular supervision may well be welcomed by those providing support, as well as risk management procedures should they feel overwhelmed by responsibilities and clinical risk scenarios. All those who take part in a peer support group may at times feel overwhelmed, and it may be important to consider ground rules on taking a break from the group or accessing alternative sources of support. Improving the confidence of people who have self-harmed to support vulnerable others might be achieved through interventions such as Mental Health First Aid Training to improve mental health literacy [31]. Peer support services might also consider providing a set of guidelines on how peers can best support others, including how to maintain one’s own emotional health in order to be in the best place to help others. More generally it will also be important when implementing peer support to provide clear signposting to other sources of support, should these be indicated alongside engagement with a peer community.

The important role that peer support plays in the lives of some people who have self-harmed suggests that this is an important dimension of a clinical assessment, and that clinicians should inquire routinely about peer support when taking a clinical history from a person who self-harms, particularly that gained online [32]. Given the acceptability of peer support to people who have self-harmed, clinicians should also gain a familiarity with the services available and discuss these as part of care planning alongside a consideration of the potential risks described. This is particularly important during a period of pandemic restrictions, when access to a full range of support sources may be severely limited. There may also be a role for clinicians in supporting the moderators or facilitators of peer support, given the importance of risk protocols and risk management when offering this type of help to a vulnerable group.

## Conclusions

Our review of the literature suggests that peer support for self-harm plays an important therapeutic role in the lives of some people who have self-harmed, who describe social, emotional and clinical benefits, but who are also able to recognise the potential for harms. This literature provides valuable suggestions for how to best implement peer support for self-harm, whether face-to-face or online, and the risk issues that that need to be considered in order to provide this safely. In view of the preferences of young people for self-harm support outside formal healthcare settings, peer support for self-harm could be a very valuable means of containing some of the distress and loneliness associated with self-harm and promoting a sense of autonomy and community.

## Lived Experience Commentary

The significance of providing peer support for those who self-harm is evidential; the current review summarises that, although not without its risks, both face-to-face and online mediums of peer support can help some young people manage self-injurious thoughts and behaviours. Through inviting open discussions around self-harm via in-person or virtual support groups, young people have reported a decrease in perceived stigma and shame compared to interventions lead by professionals or loved ones. This may be attributed to a sense of solidarity, whereby those who self-harm are provided with a safe space to share their narratives and support each others’ struggles and goals, during times where they may feel isolated and misunderstood by those who do not share similar experiences.

From my perspective, this review has brought to light how society has altered the way young people choose to obtain informal support. Although face-to-face groups hold their advantages, it is interesting to note that online platforms are an increasingly popular means for seeking help, due to giving/receiving support from a range of like-minded people, and the instant access to useful information. These findings strongly resonate with my own lived experience as an adolescent; I found solace in the mutual benefits of sharing emotional distress and self-harm ideation with others, whilst maintaining anonymity behind a mobile screen. However, the associated risks of using online mediums should not go unnoticed; including exposure to graphic images of extreme self-harm and feeling overwhelmed with a perceived responsibility to support others.

Although I understand the high-risk nature of peer support in increasing some self-harm behaviours, I view the benefits to outweigh the cons in terms of accessing support and generating a sense of self-empowerment and online community presence for young people. Given these potential benefits, and in light of the few studies investigating its effectiveness of peer interventions, there is an urgent need to determine its efficacy as an active intervention. Only through clinical trials can risk issues concerning both group members and facilitators/moderators be fully addressed, and peer interventions could start to be considered with the same necessity for managing self-harm as professional support services.

## Data Availability

An example of our search strategy is provided in the supplementary material

## Abbreviations PRISMA

UK: United Kingdom
GRADE: Grading of Recommendations, Assessment, Development, and Evaluation
CASP: Critical Appraisals Skill Programme
MMAT: Mixed Methods Appraisal Tool
NESTA: NESTA’s Standard of Evidence model
US: United States

## 3. Studies and reports included in the main results

**Table 1A:**
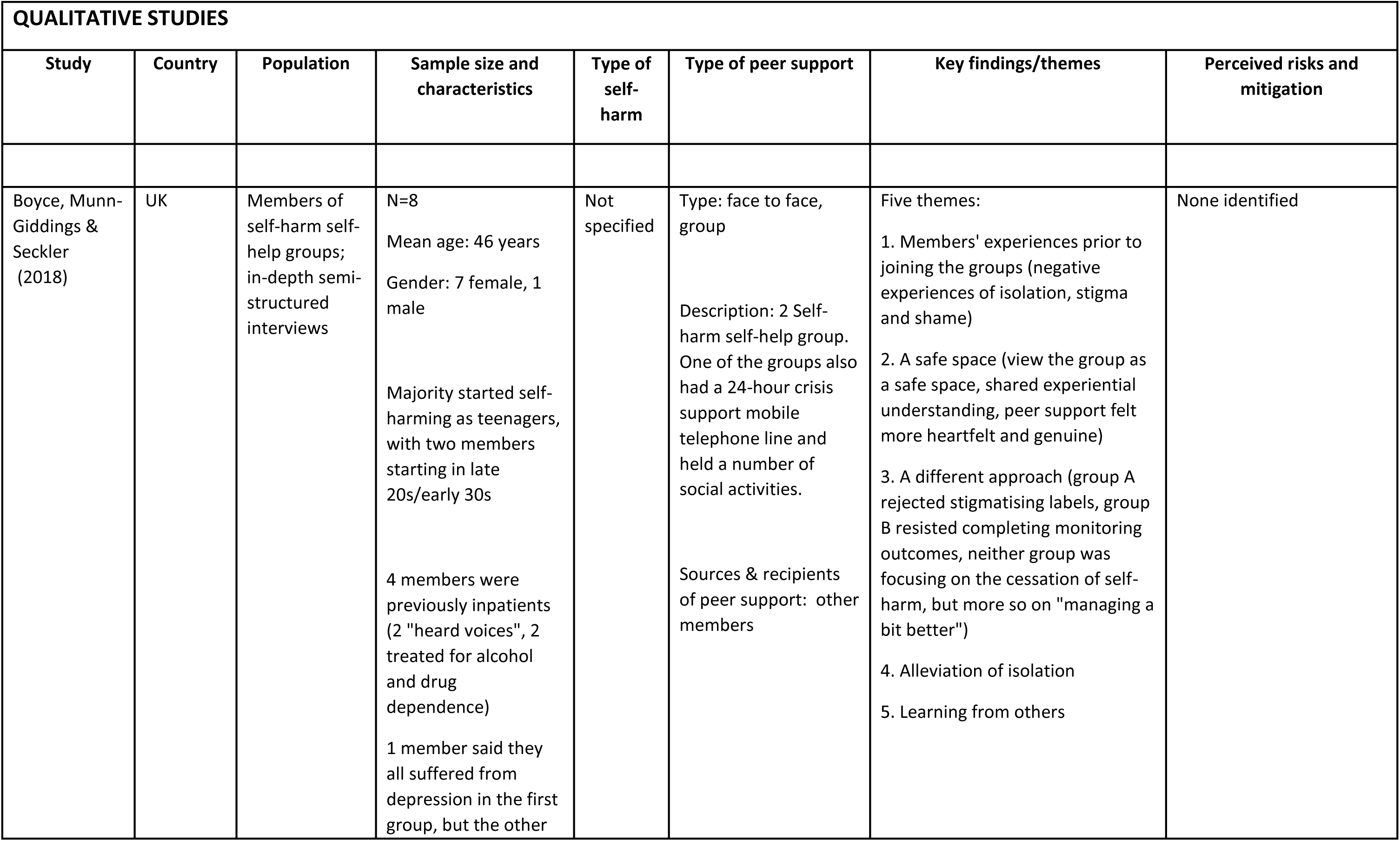
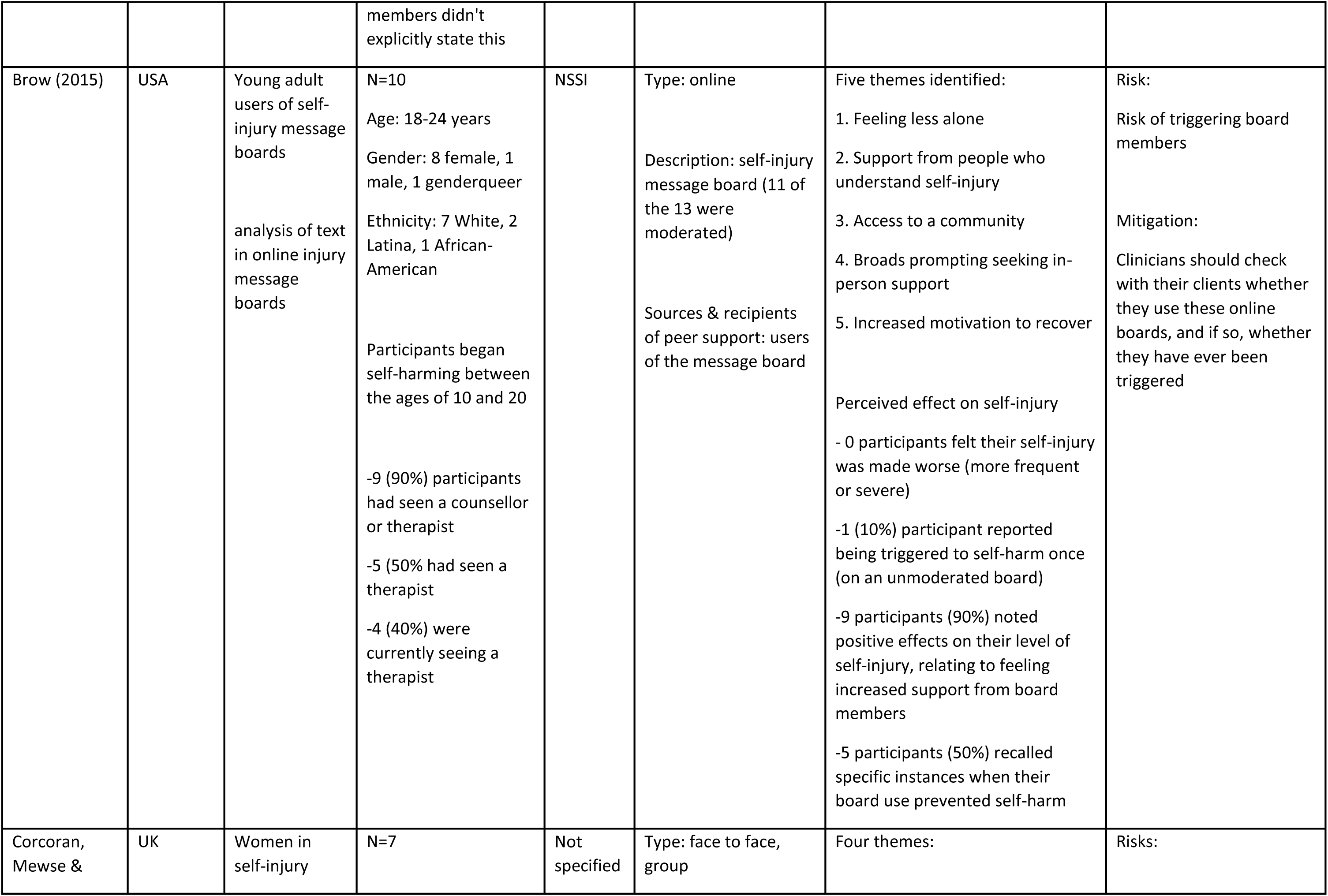
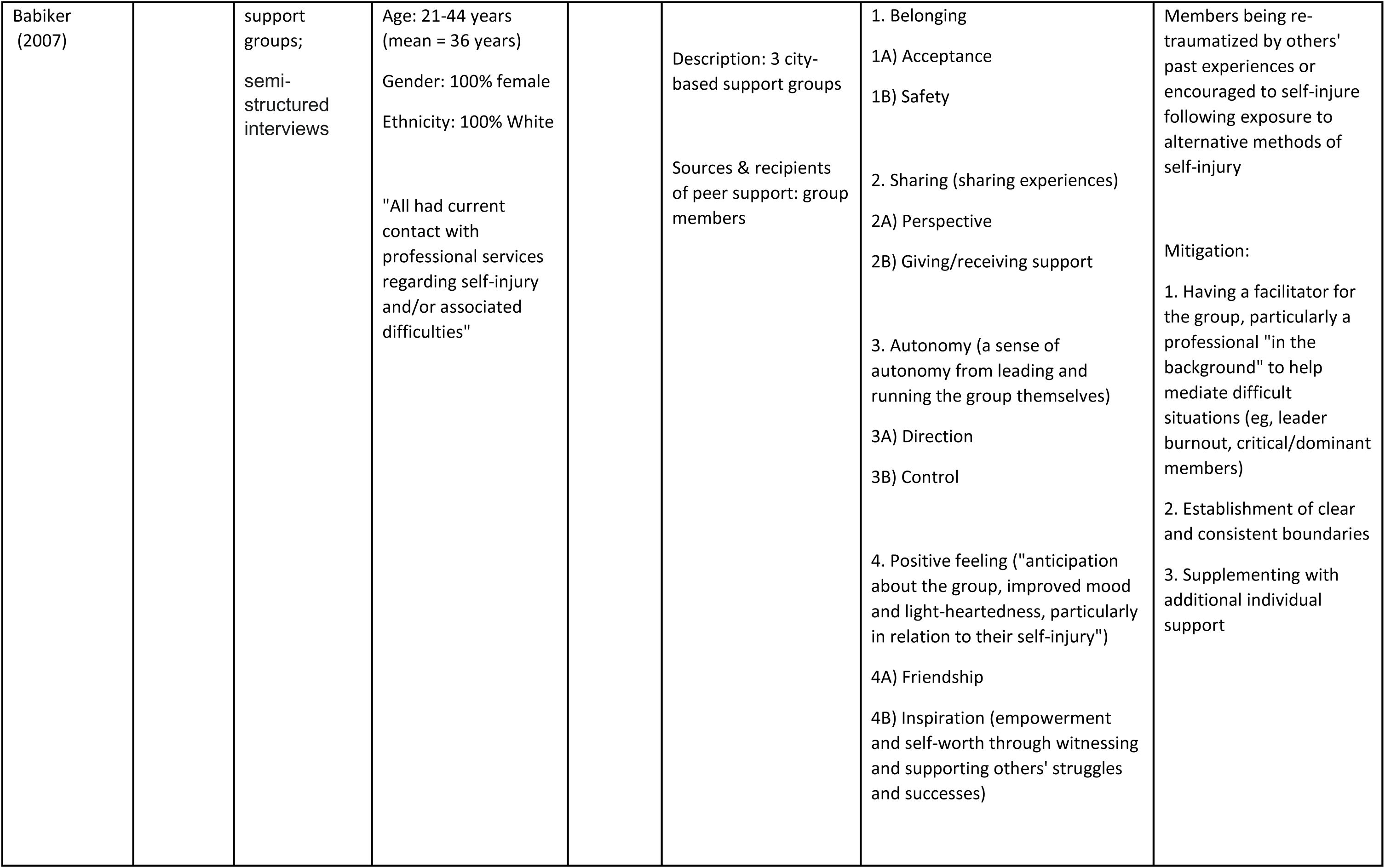
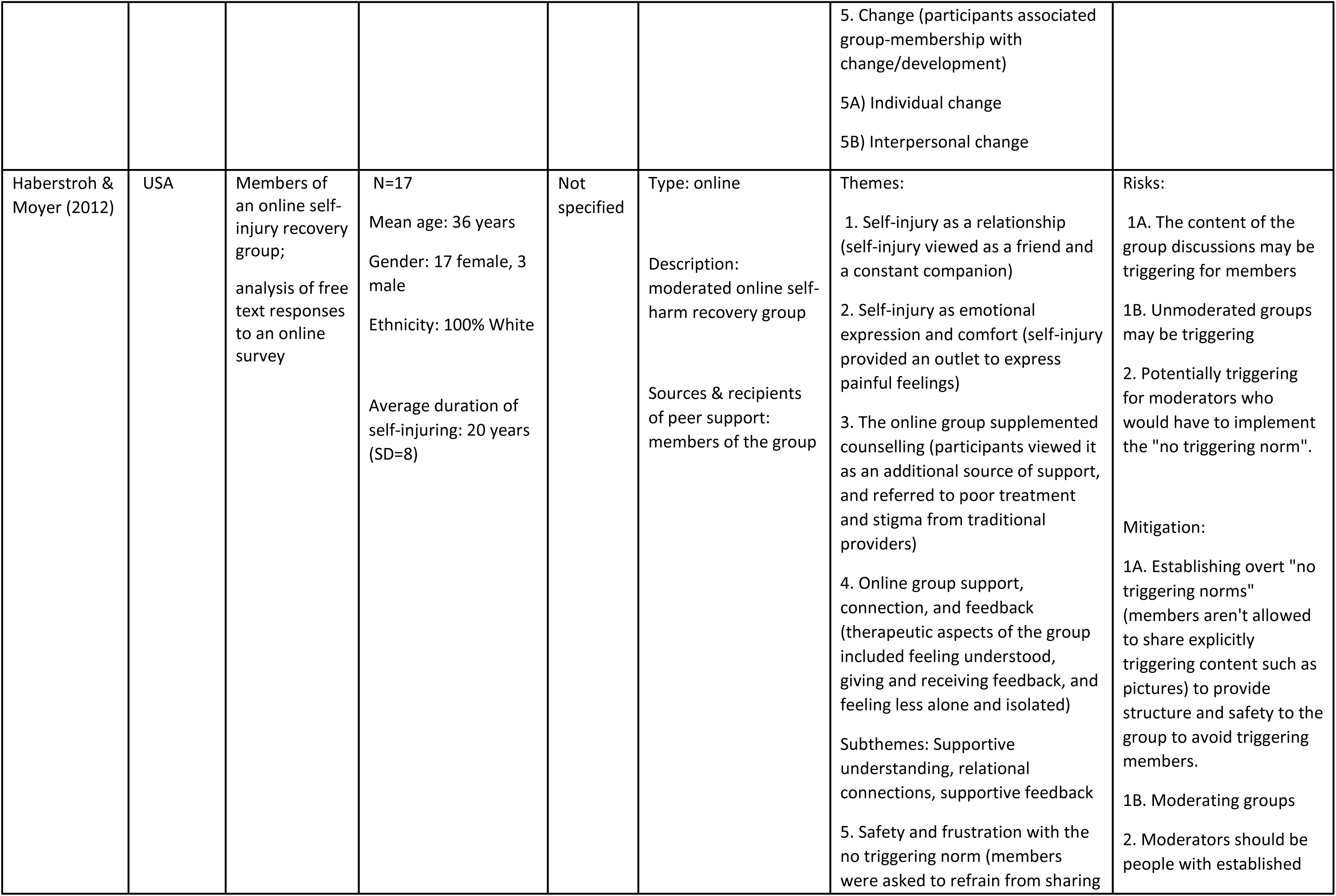
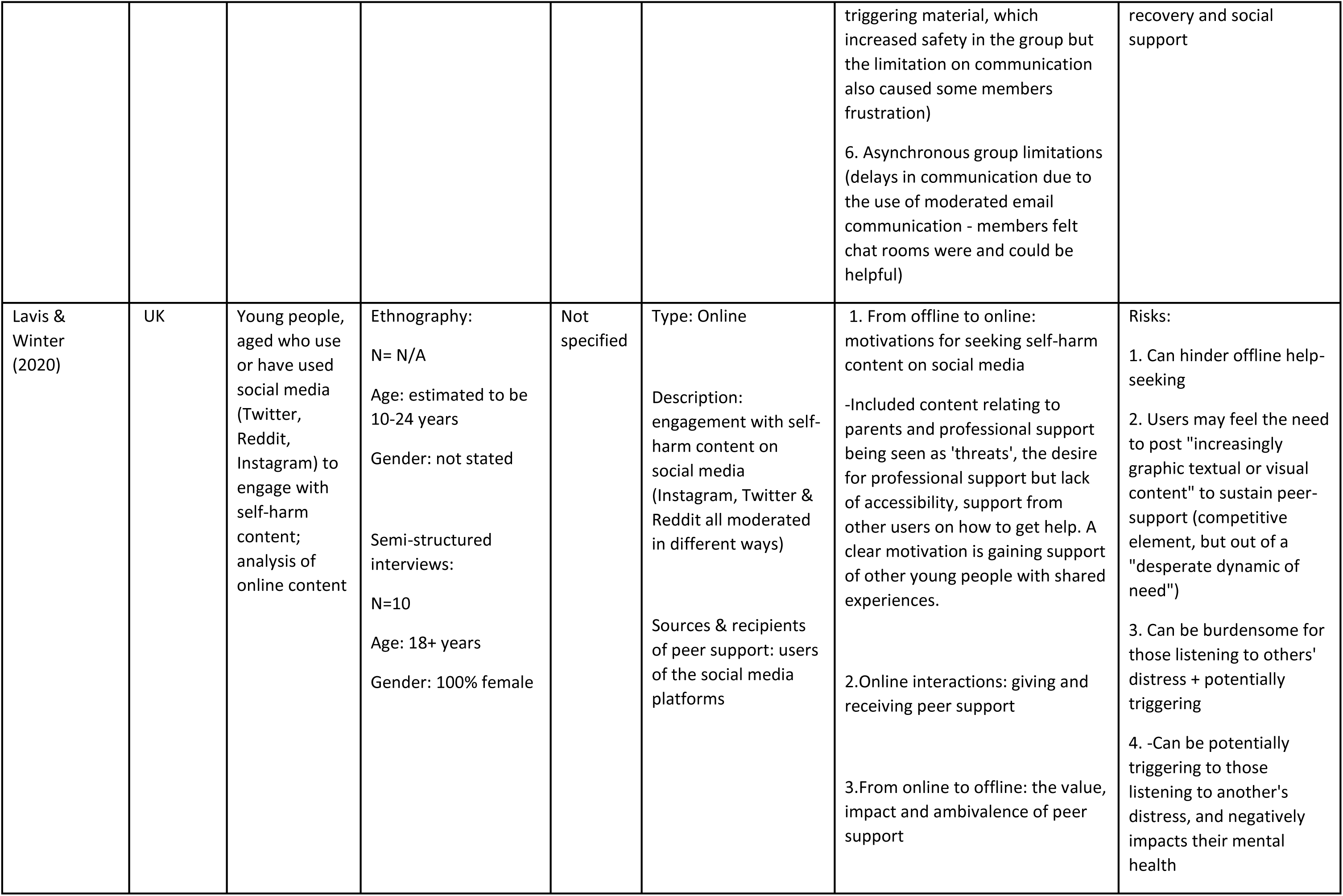
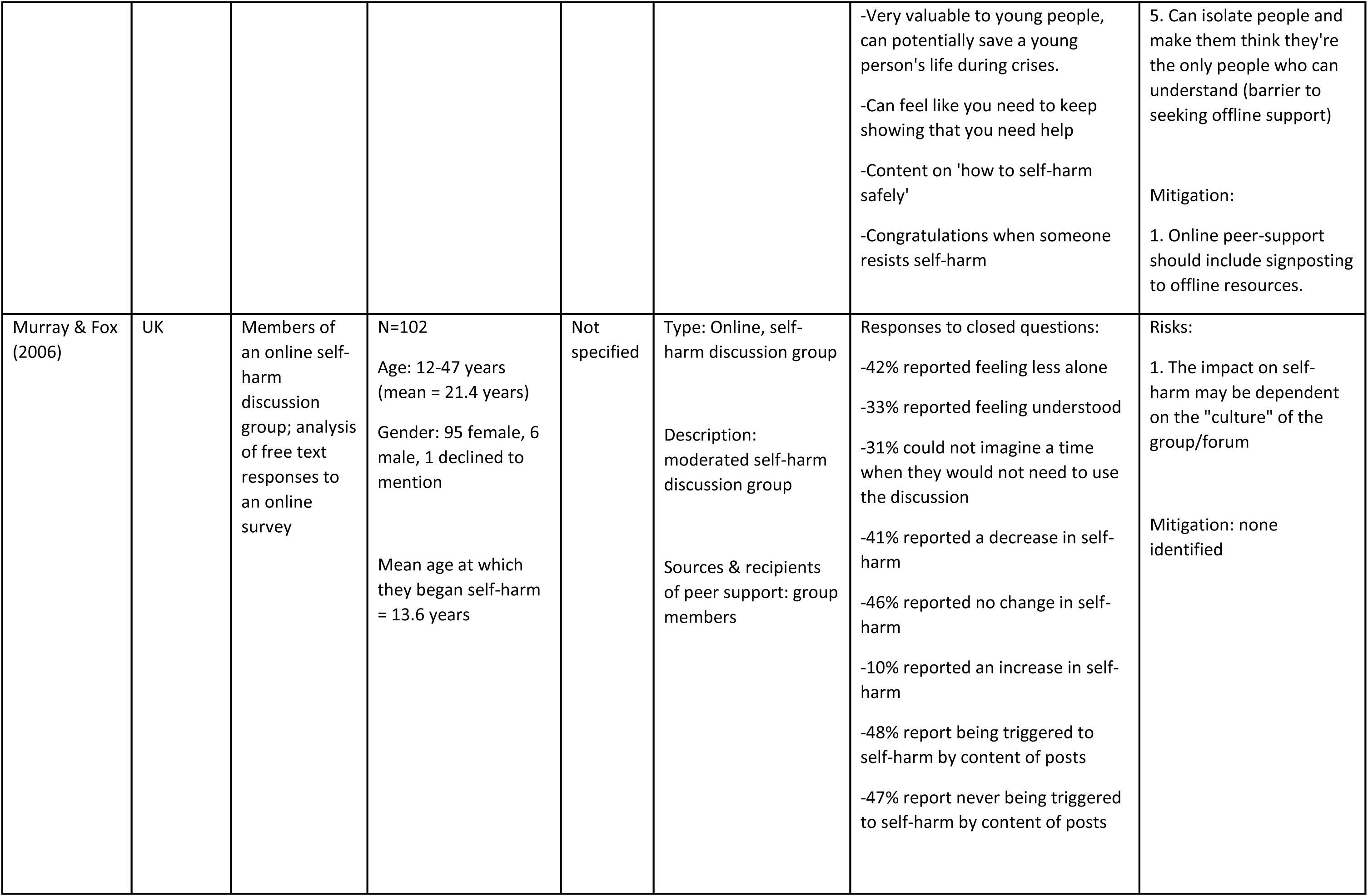
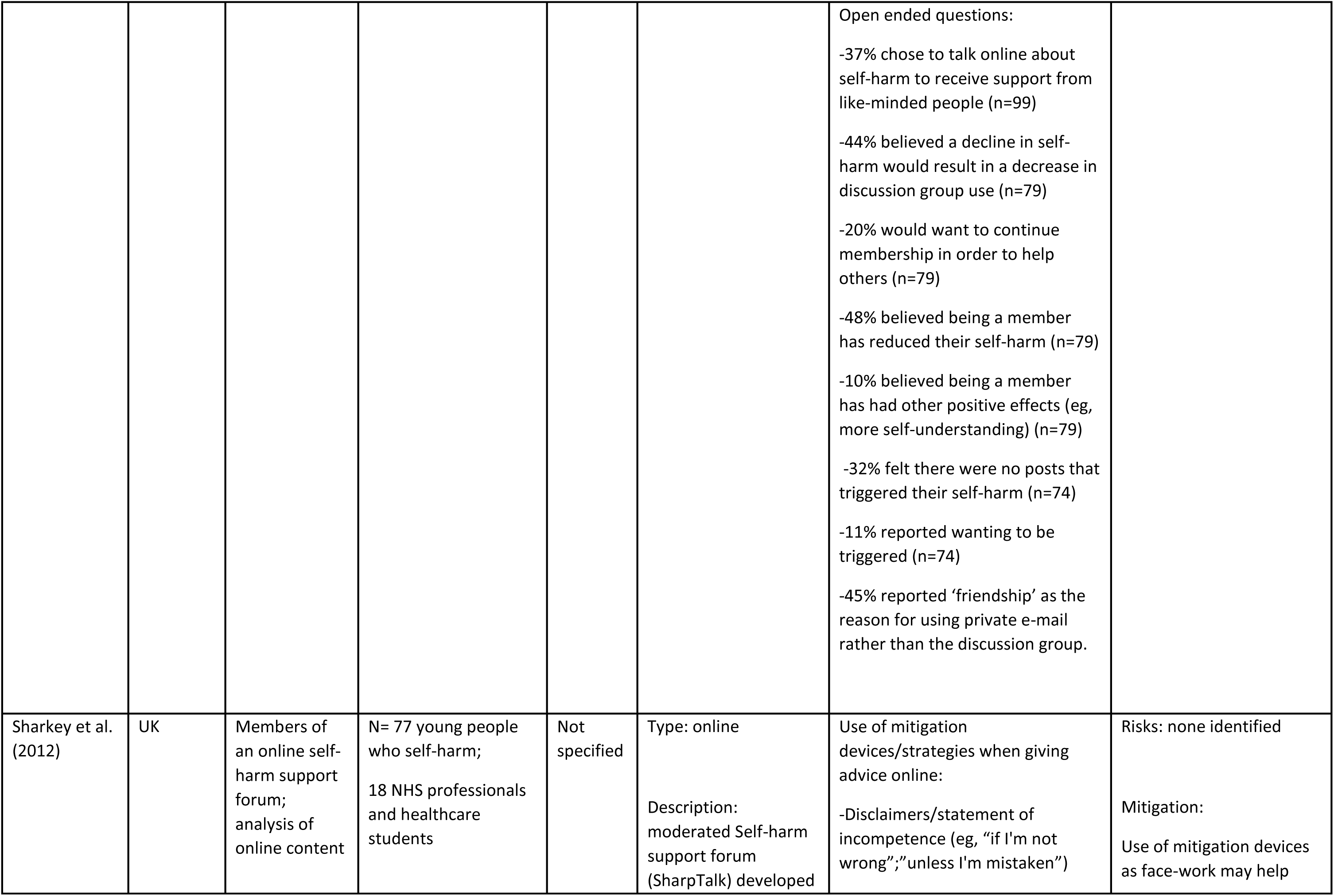
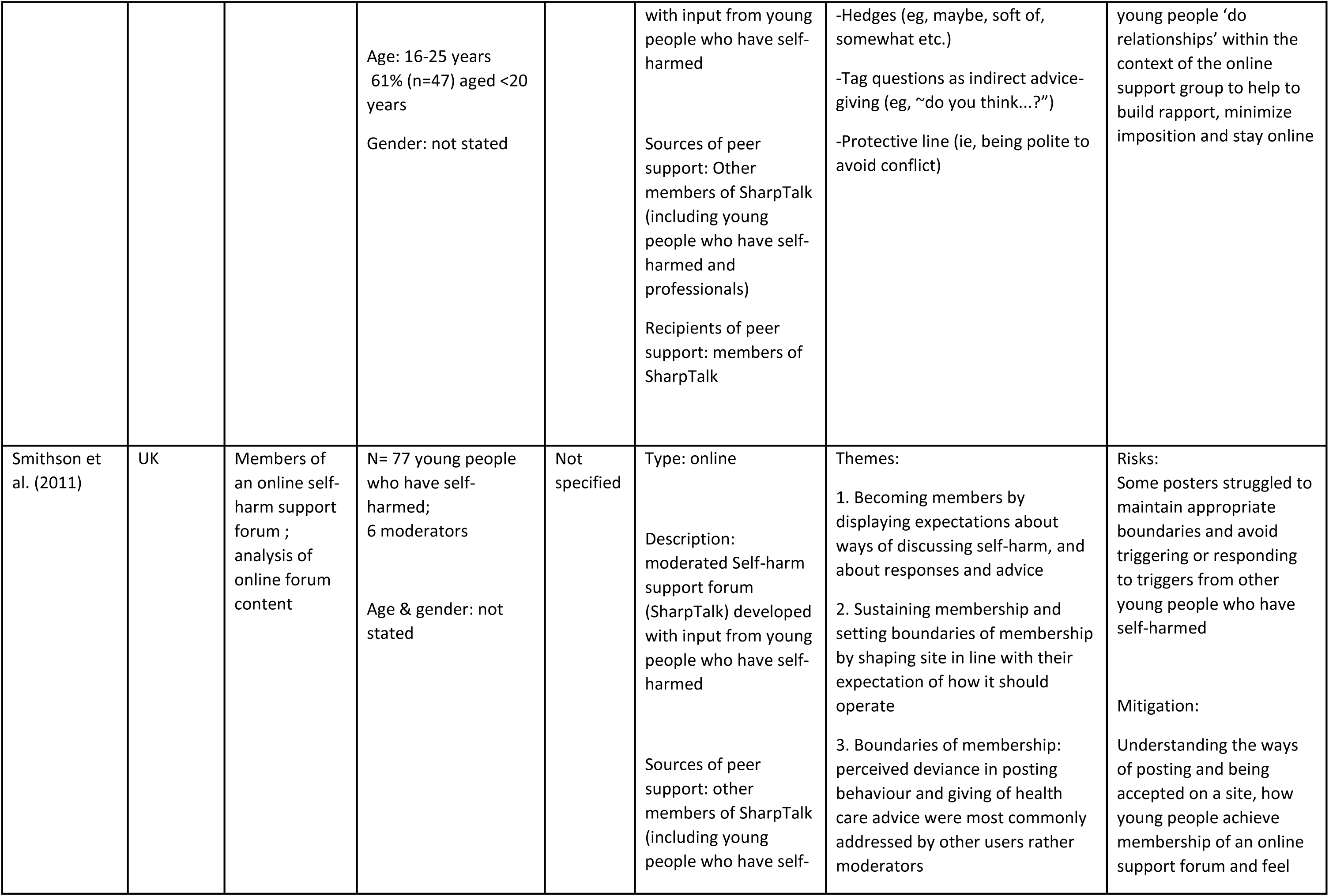
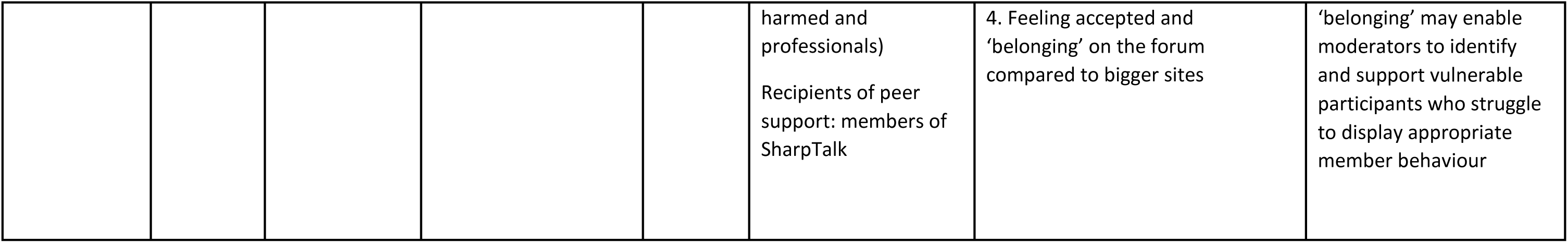
Qualitative studies and reports included in main results

**Table 1B:**
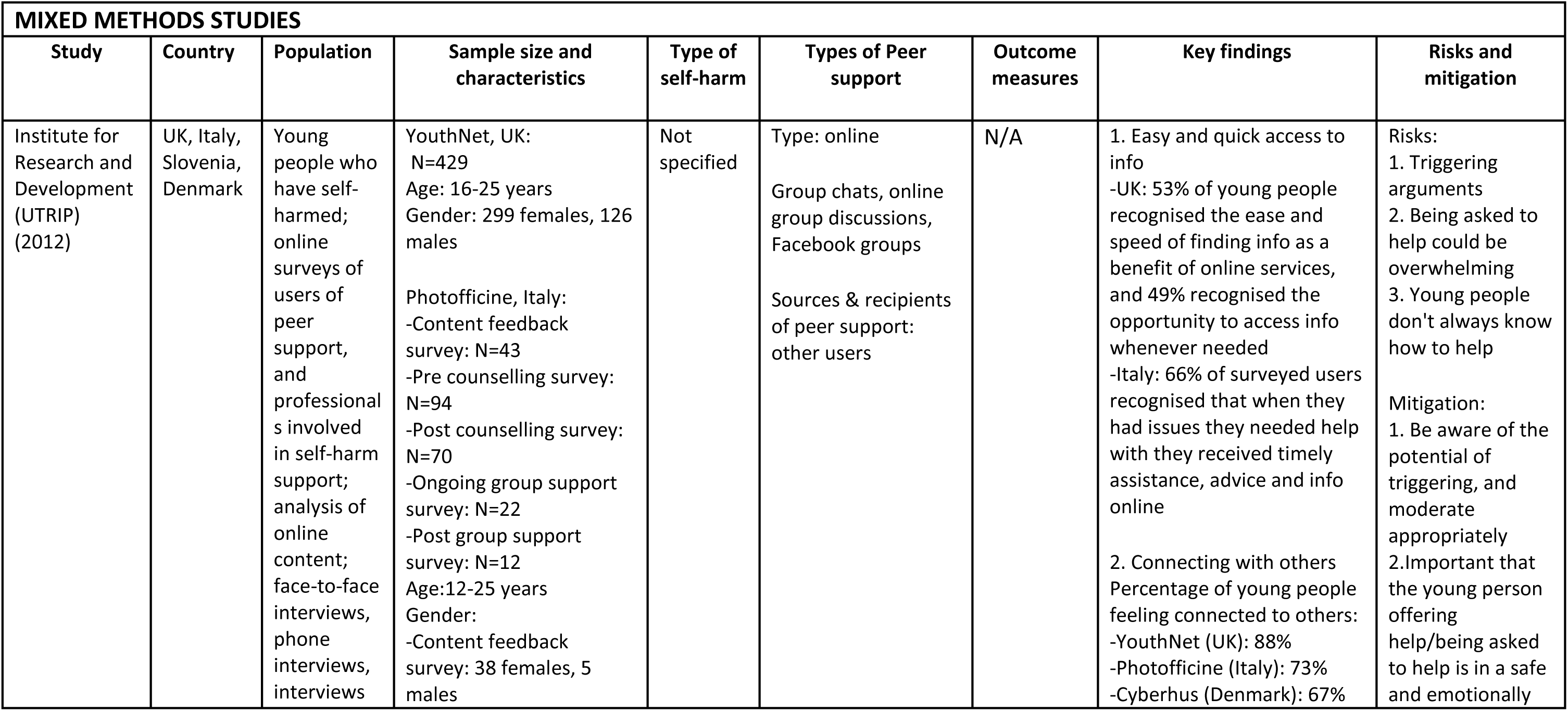
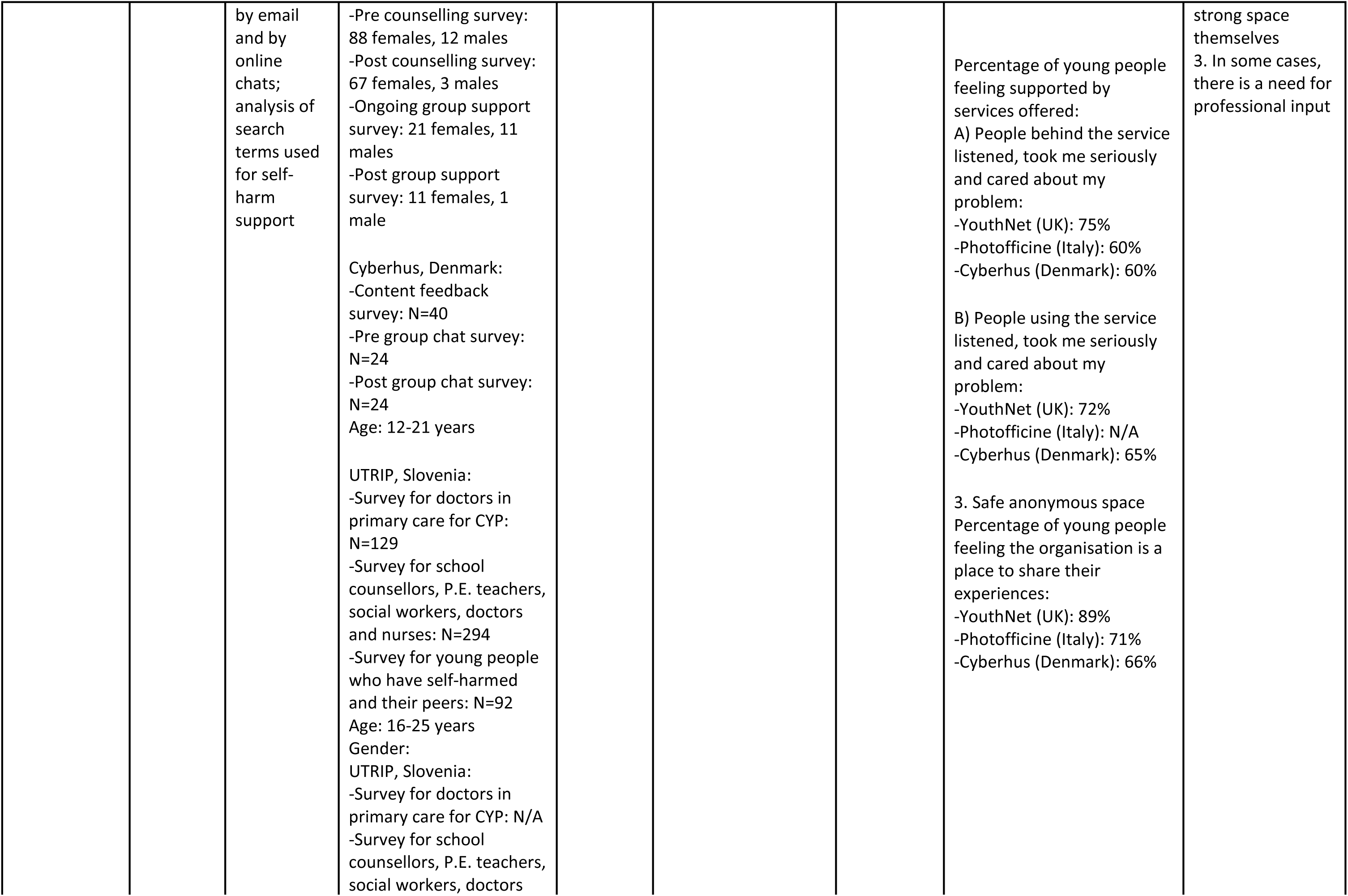
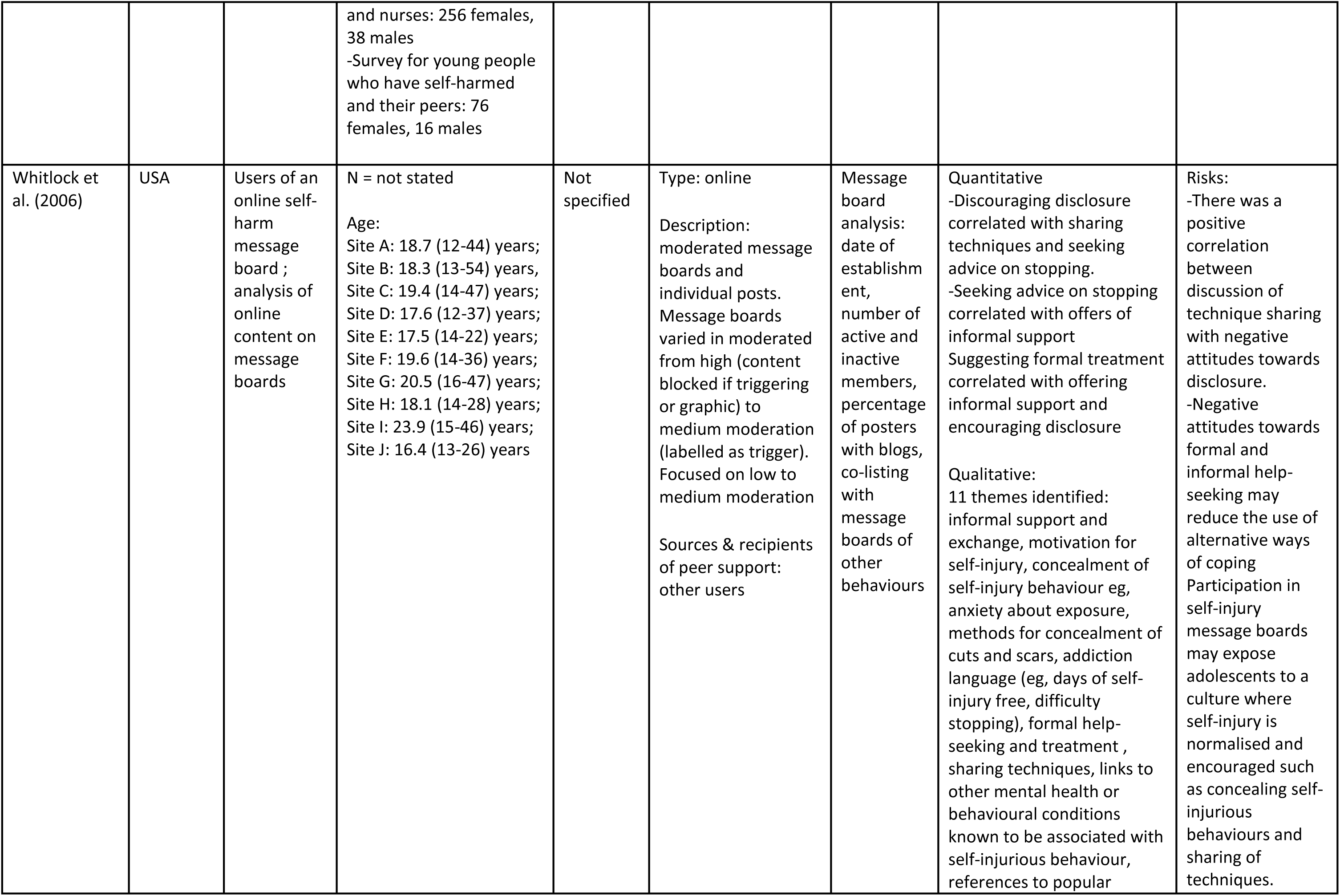
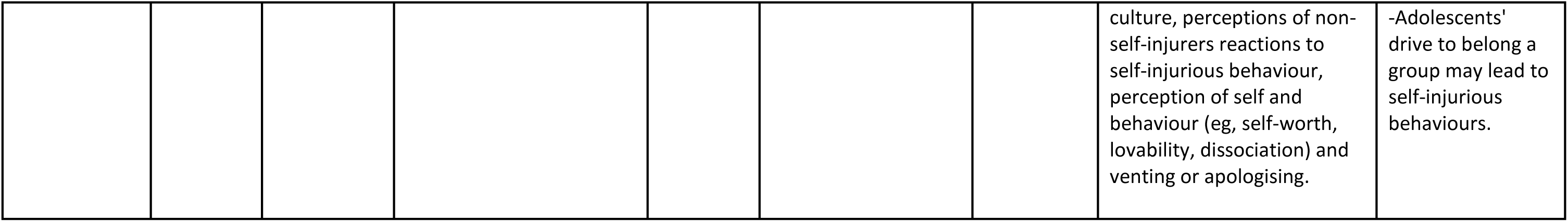
Mixed methods studies and reports included in main results

## Supplementary material

### 1. Search Strategies

#### 1A. MEDLINE

1. self-injurious behavior/ or self mutilation/ (11249)
2. (parasuicide or “suicidal self-injur” or “suicidal self?injur*” or SSI or “self-injurious behavi*” or “self?injurious behavi*” or “suicidal behavi*” or automutilation or auto-mutilation or “auto?mutilation” or self-mutilation or “self?mutilation” or “self-inflicted wound*” or “self?inflicted wound*” or “self-inflicted injur*” or “self?inflicted injur*” or “non-suicidal self-injur*” or “non-suicidal self? injur*” or NSSI).tw. (21766)
3. (self-harm* or “self?harm*” or self-injur* or “self?injur*” or self-destruct* or “self?destruct*” or self-poison* or “self?poison*” or self-immolat* or “self?immolat*” or self-inflict* or “self?inflict*” or auto-destruct* or “auto?destruct*” or (self adj2 cut*) or cut*).tw. (439664)
4. 1 or 2 or 3 (459740)
5. peer group/ or Social support/ or Self-help groups/ or Education, Nonprofessional/ or Psychosocial support systems/ or Community networks/ (105183)
6. (((peer* or support* or volunteer* or “lived experience” or “service user*” or community or lay or mutual) adj2 (group* or network* or communit* or relation* or support* or listen* or help* or visit* or aid*)) or ((social or community) adj3 (support or network* or group*)) or ((peer* or lay or volunteer *) adj (based or counsel* or deliver* or interact* or led or mediat* or operat* or provid* or run*)) or “peer support” or peer-to-peer or “peer?to?peer” or peer* or “social interaction program*” or befriend*).tw. (2147753)
7. (((lay or peer) adj2 (advisor* or consultant* or educator* or expert* or facilitator* or instructor* or leader* or person* or tutor* or worker* or advice* or advise* or counsel* or mentor*)) or “expert patient*”).tw. (6624)
8. 5 or 6 or 7 (2181210)
9. (”facebook” or e-mail or e-support or “e-bulletin board” or e-society or e-community or forum or “discussion group” or “message board” or “internet relay” or “instant?messaging” or “chat technology” or “chat room” or chatroom or chatgroup or “chat group” or ((virtual or internet or online or web-based or “web?based” or internet-based or “internet?based”) adj2 (support or network or group or society or community or interaction or “bulletin board”))).tw. (30668)
10. (helpline or “help?line” or ((phone* or telephone*) adj3 (help* or instruct* or interact* or mediat* or program* or rehab* or strateg* or support* or teach* or train* or workshop* or based or assist* or driven or led))).tw. (11366)
11. 9 or 10 (41824)
12. 1 (”peer?support” or peer-support or “self?help” or self-help or “social support” or “support group” or “support network” or “support system”).tw. (62443)
13. 11 and 12 (2371)
14. 8 or 13 (2181336)
15. 4 and 14 (36840)
16. limit 15 to (english language and humans and yr=”2000 -Current”) (19899)

#### 1B. PsycINFO

1. self-injurious behavior/ or self mutilation/
2. (parasuicide or “suicidal self-injur” or “suicidal self?injur*” or SSI or “self-injurious behavi*” or “self?injurious behavi*” or “suicidal behavi*” or automutilation or auto-mutilation or “auto?mutilation” or self-mutilation or “self?mutilation” or “self-inflicted wound*” or “self?inflicted wound*” or “self-inflicted injur*” or “self?inflicted injur*” or “non-suicidal self-injur*” or “non-suicidal self? injur*” or NSSI).tw. (17882)
3. (self-harm* or “self?harm*” or self-injur* or “self?injur*” or self-destruct* or “self?destruct*” or self-poison* or “self?poison*” or self-immolat* or “self?immolat*” or self-inflict* or “self?inflict*” or auto-destruct* or “auto?destruct*” or (self adj2 cut*) or cut*).tw. (58008)
4. 1 or 2 or 3 (69788)
5. peer group/ or Social support/ or Self-help groups/ or Education, Nonprofessional/ or Psychosocial support systems/ or Community networks/ (37486)
6. (((peer* or support* or volunteer* or “lived experience” or “service user*” or community or lay or mutual) adj2 (group* or network* or communit* or relation* or support* or listen* or help* or visit* or aid*)) or ((social or community) adj3 (support or network* or group*)) or ((peer* or lay or volunteer *) adj (based or counsel* or deliver* or interact* or led or mediat* or operat* or provid* or run*)) or “peer support” or peer-to-peer or “peer?to?peer” or peer* or “social interaction program*” or befriend*).tw. (998301)
7. (((lay or peer) adj2 (advisor* or consultant* or educator* or expert* or facilitator* or instructor* or leader* or person* or tutor* or worker* or advice* or advise* or counsel* or mentor*)) or “expert patient*”).tw. (7396)
8. 5 or 6 or 7 (1000384)
9. (”facebook” or e-mail or e-support or “e-bulletin board” or e-society or e-community or forum or “discussion group” or “message board” or “internet relay” or “instant?messaging” or “chat technology” or “chat room” or chatroom or chatgroup or “chat group” or ((virtual or internet or online or web-based or “web?based” or internet-based or “internet?based”) adj2 (support or network or group or society or community or interaction or “bulletin board”))).tw. (24369)
10. (helpline or “help?line” or ((phone* or telephone*) adj3 (help* or instruct* or interact* or mediat* or program* or rehab* or strateg* or support* or teach* or train* or workshop* or based or assist* or driven or led))).tw. (6334)
11. 9 or 10 (30591)
12. (”peer?support” or peer-support or “self?help” or self-help or “social support” or “support group” or “support network” or “support system”).tw. (71602)
13. 11 and 12 (2310)
14. 8 or 13 (1000529)
15. 4 and 14 (15315)
16. limit 15 to (human and english language and yr=”2000 -Current”) (11768)

#### 1C. Open Grey

▪ Self injury and social support
▪ self injury and self help groups
▪ self injury and psychosocial support
▪ self injury and community networks
▪ self harm AND peer group
▪ self harm AND social support
▪ Self harm and self help group
▪ Self harm and lived experience
▪ self harm and volunteer
▪ Self harm and lay expert
▪ suicidal self injury and peer support
▪ suicidal self injury and social support
▪ non-suicidal self injury and social support
▪ non-suicidal self injury and peer support
▪ NSSI and social support
▪ NSSI and peer support
▪ parasuicide and support group
▪ parasuicide and peer group
▪ parasuicide and peer support
▪ self injury and volunteer
▪ self injury and lay expert
▪ Self mutilation and peer group
▪ Self injurious and ‘peer group’

### 2. Websites searched and organisations contacted for grey literature search

- We Are with You (formerly known as Addaction)
- Mind
- Mental Health Foundation
- Rethink Mental Illness
- Time to Change
- Together for Mental Wellbeing
- Harmless
- Young Minds
- Agenda
- Beat
- Anorexia, Bulimia and Care (ABC)
- Overeaters Anonymous
- Alcoholics Anonymous
- Narcotics Anonymous
- Frank

### 3. Quality Appraisal

**Table 3A:**
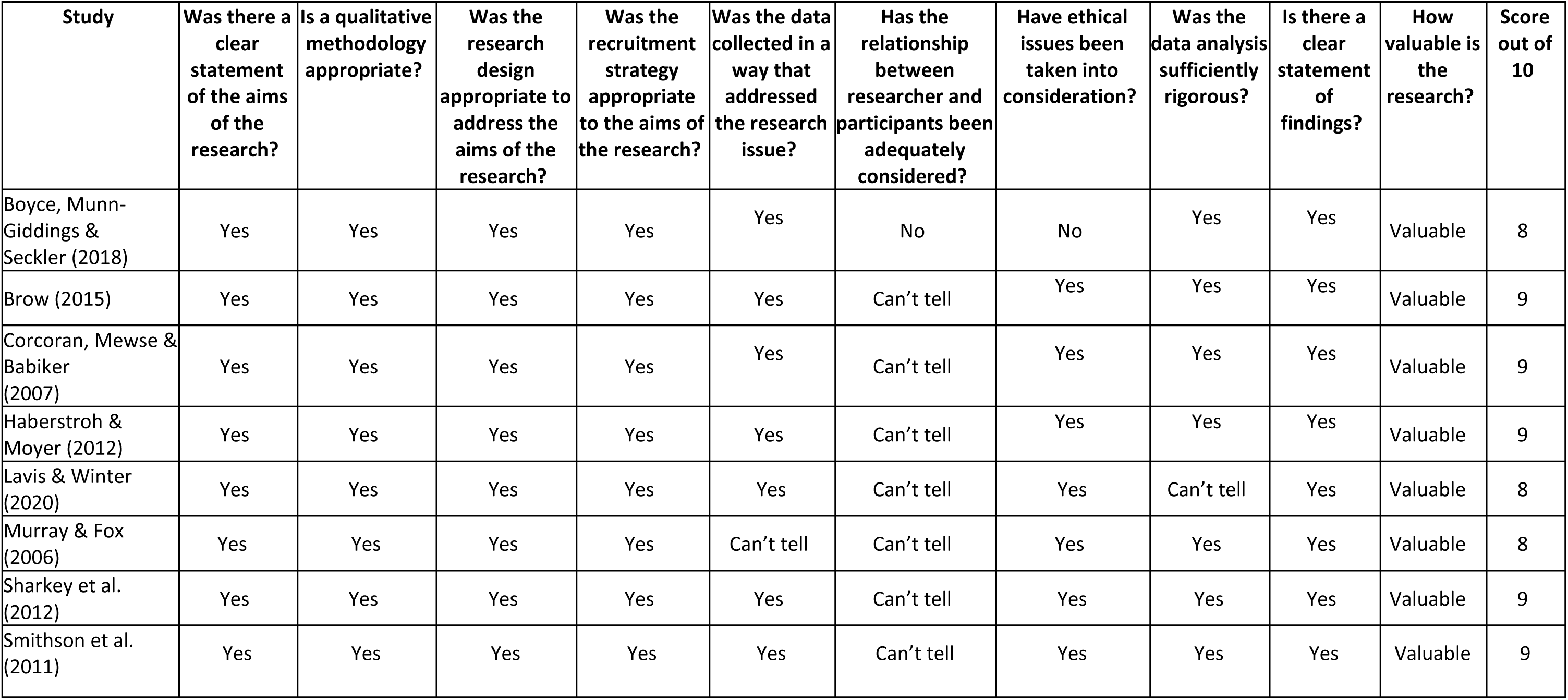
Quality appraisal of qualitative studies using the Critical Appraisals Skill Programme (CASP) Qualitative Checklist

**Table 3B:** Quality appraisal of mixed methods studies using the Mixed Methods Appraisal Tool (MMAT)

| Study | Qualitative |  |  |  |  | Quantitative |  |  |  |  | Mixed Methods |  |  |  |  | Score out of 15 |
| --- | --- | --- | --- | --- | --- | --- | --- | --- | --- | --- | --- | --- | --- | --- | --- | --- |
|  | 1.1 | 1.2 | 1.3 | 1.4 | 1.5 | 4.1 | 4.2 | 4.3 | 4.4 | 4.5 | 5.1 | 5.2 | 5.3 | 5.4 | 5.5 |  |
| Whitlock et al. (2006) | Yes | Yes | Yes | Yes | Yes | Yes | Can't tell | Can't tell | Can't tell | Yes | Yes | Yes | Yes | Yes | Can't tell | 11 |

**Table 3C:** Quality appraisal of reports not presented as primary research studies using NESTA’s Standards of Evidence

| Study | Level 1: Have they described what positive difference this innovation is going to make? | Level 2: Is there data that shows the positive difference can be measured? | Level 3: Have they proved that the positive difference is because of the innovation (by testing with comparison groups)? | Level 4: Have other people proved the same thing? | Level 5: Can this innovation be replicated elsewhere? | Levels achieved |
| --- | --- | --- | --- | --- | --- | --- |
| Institute for Research and Development "UTRIP" (2012) | Yes | Yes | Not sure | Yes | Yes | 2 |

| Section and Topic | Item # | Checklist item | Location where item is reported |
| --- | --- | --- | --- |
| **TITLE** |  |  |  |
| Title | 1 | Identify the report as a systematic review. | 1 |
| **ABSTRACT** |  |  |  |
| Abstract | 2 | See the PRISMA 2020 for Abstracts checklist. | 2-3 |
| **INTRODUCTION** |  |  |  |
| Rationale | 3 | Describe the rationale for the review in the context of existing knowledge. | 4 |
| Objectives | 4 | Provide an explicit statement of the objective(s) or question(s) the review addresses. | 5 |
| **METHODS** |  |  |  |
| Eligibility criteria | 5 | Specify the inclusion and exclusion criteria for the review and how studies were grouped for the syntheses. | 7-9 |
| Information sources | 6 | Specify all databases, registers, websites, organisations, reference lists and other sources searched or consulted to identify studies. Specify the date when each source was last searched or consulted. | 6 |
| Search strategy | 7 | Present the full search strategies for all databases, registers and websites, including any filters and limits used. | 47-50 |
| Selection process | 8 | Specify the methods used to decide whether a study met the inclusion criteria of the review, including how many reviewers screened each record and each report retrieved, whether they worked independently, and if applicable, details of automation tools used in the process. | 8 |
| Data collection process | 9 | Specify the methods used to collect data from reports, including how many reviewers collected data from each report, whether they worked independently, any processes for obtaining or confirming data from study investigators, and if applicable, details of automation tools used in the process. | 8-9 |
| Data items | 10a | List and define all outcomes for which data were sought. Specify whether all results that were compatible with each outcome domain in each study were sought (e.g. for all measures, time points, analyses), and if not, the methods used to decide which results to collect. | 9 |
|  | 10b | List and define all other variables for which data were sought (e.g. participant and intervention characteristics, funding sources). Describe any assumptions made about any missing or unclear information. | 8-9 |
| Study risk of bias assessment | 11 | Specify the methods used to assess risk of bias in the included studies, including details of the tool(s) used, how many reviewers assessed each study and whether they worked independently, and if applicable, details of automation tools used in the process. | 9-10 |
| Effect measures | 12 | Specify for each outcome the effect measure(s) (e.g. risk ratio, mean difference) used in the synthesis or presentation of results. |  |
| Synthesis methods | 13a | Describe the processes used to decide which studies were eligible for each synthesis (e.g. tabulating the study intervention characteristics and comparing against the planned groups for each synthesis (item #5)). | 9-10 |
|  | 13b | Describe any methods required to prepare the data for presentation or synthesis, such as handling of missing summary statistics, or data conversions. | 9-10 |
|  | 13c | Describe any methods used to tabulate or visually display results of individual studies and syntheses. |  |
|  | 13d | Describe any methods used to synthesize results and provide a rationale for the choice(s). If meta-analysis was performed, describe the model(s), method(s) to identify the presence and extent of statistical heterogeneity, and software package(s) used. | n/a |
|  | 13e | Describe any methods used to explore possible causes of heterogeneity among study results (e.g. subgroup analysis, meta-regression). | n/a |
|  | 13f | Describe any sensitivity analyses conducted to assess robustness of the synthesized results. | n/a |
| Reporting bias assessment | 14 | Describe any methods used to assess risk of bias due to missing results in a synthesis (arising from reporting biases). |  |
| Certainty assessment | 15 | Describe any methods used to assess certainty (or confidence) in the body of evidence for an outcome. |  |
| **RESULTS** |  |  |  |
| Study selection | 16a | Describe the results of the search and selection process, from the number of records identified in the search to the number of studies included in the review, ideally using a flow diagram. | 12 |
|  | 16b | Cite studies that might appear to meet the inclusion criteria, but which were excluded, and explain why they were excluded. |  |
| Study characteristics | 17 | Cite each included study and present its characteristics. | 34-45 |
| Risk of bias in studies | 18 | Present assessments of risk of bias for each included study. | 51-52 |
| Results of individual studies | 19 | For all outcomes, present, for each study: (a) summary statistics for each group (where appropriate) and (b) an effect estimate and its precision (e.g. confidence/credible interval), ideally using structured tables or plots. | n/a |
| Results of syntheses | 20a | For each synthesis, briefly summarise the characteristics and risk of bias among contributing studies. | 14-18 |
|  | 20b | Present results of all statistical syntheses conducted. If meta-analysis was done, present for each the summary estimate and its precision (e.g. confidence/credible interval) and measures of statistical heterogeneity. If comparing groups, describe the direction of the effect. | n/a |
|  | 20c | Present results of all investigations of possible causes of heterogeneity among study results. | n/a |
|  | 20d | Present results of all sensitivity analyses conducted to assess the robustness of the synthesized results. | n/a |
| Reporting biases | 21 | Present assessments of risk of bias due to missing results (arising from reporting biases) for each synthesis assessed. | n/a |
| Certainty of evidence | 22 | Present assessments of certainty (or confidence) in the body of evidence for each outcome assessed. | 19-20 |
| **DISCUSSION** |  |  |  |
| Discussion | 23a | Provide a general interpretation of the results in the context of other evidence. |  |
|  | 23b | Discuss any limitations of the evidence included in the review. | 22-24 |
|  | 23c | Discuss any limitations of the review processes used. | 22-24 |
|  | 23d | Discuss implications of the results for practice, policy, and future research. | 24-26 |
| **OTHER INFORMATION** |  |  |  |
| Registration and protocol | 24a | Provide registration information for the review, including register name and registration number, or state that the review was not registered. |  |
|  | 24b | Indicate where the review protocol can be accessed, or state that a protocol was not prepared. | 5 |
|  | 24c | Describe and explain any amendments to information provided at registration or in the protocol. |  |
| Support | 25 | Describe sources of financial or non-financial support for the review, and the role of the funders or sponsors in the review. | 30 |
| Competing interests | 26 | Declare any competing interests of review authors. | 30 |
| Availability of data, code and other materials | 27 | Report which of the following are publicly available and where they can be found: template data collection forms; data extracted from included studies; data used for all analyses; analytic code; any other materials used in the review. |  |

## References

1. Madge, N., et al., Deliberate self-harm within an international community sample of young people: comparative findings from the Child & Adolescent Self-harm in Europe (CASE) Study. Journal of child psychology and psychiatry, and allied disciplines, 2008. 49(6): p. 667–77.

2. Muehlenkamp, J.J. and P.M. Gutierrez, An Investigation of Differences Between Self-Injurious Behavior and Suicide Attempts in a Sample of Adolescents. Suicide and Life-Threatening Behavior, 2004. 34(1): p. 12–23.

3. Hiscock, H., et al., Paediatric mental and physical health presentations to emergency departments, Victoria, 2008-15. Med J Aust, 2018. 208(8): p. 343–348.

4. McManus, S., et al., Prevalence of non-suicidal self-harm and service contact in England, 2000-14: repeated cross-sectional surveys of the general population. The lancet. Psychiatry, 2019. 6(7): p. 573–581.

5. Tormoen, A.J., et al., Change in prevalence of self-harm from 2002 to 2018 among Norwegian adolescents. Eur J Public Health, 2020. 30(4): p. 688–692.

6. Patalay, P., & Fitzsimons, E., Psychological distress, self-harm and attempted suicide in UK 17-year olds: prevalence and sociodemographic inequalities. The British Journal of Psychiatry, 2021: p. 1–3.

7. Hawton, K., et al., Deliberate self-harm in Oxford, 1990-2000: a time of change in patient characteristics. Psychological medicine, 2003. 33(6): p. 987–95.

8. Rowe, S.L., et al., Help-seeking behaviour and adolescent self-harm: a systematic review. The Australian and New Zealand journal of psychiatry, 2014. 48(12): p. 1083–95.

9. Basset, T., Faulkner, A., Repper, J. and Stamou, E., Lived experience leading the way: peer support in mental health. 2010.

10. Shalaby, R.A.H. and V.I.O. Agyapong, Peer Support in Mental Health: Literature Review. JMIR Ment Health, 2020. 7(6): p. e15572.

11. Self-Harm., A.-P.P.G.o.S.a., Inquiry into the support available for young people who self-harm. 2020.

12. Stevinson, C., Lawlor, DA., Searching multiple databases for systematic reviews: added value or diminishing returns? Complement Ther Med, 2004. 12(4): p. 228–32.

13. McHugh, M.L., Interrater reliability: the kappa statistic. Biochem Med (Zagreb), 2012. 22(3): p. 276–282.

14. Schünemann H, B.J., Guyatt G, Oxman A,, GRADE handbook for grading quality of evidence and strength of recommendations. Updated October 2013.

15. UK., C. Critical Appraisal Skills Programme. CASP (Qualitative Checklist) . [online]. 2018 March 2021]; Available from: https://casp-uk.net/wp-content/uploads/2018/01/CASP-Systematic-Review-Checklist_2018.pdf.

16. Hong, Q.N., Fàbregues, S., Bartlett, G., Boardman, F., Cargo, M., Dagenais, P., Gagnon, M. P., Griffiths, F., Nicolau, B., O’Cathain, A., Rousseau, M. C., Vedel, I., & Pluye, P., The Mixed Methods Appraisal Tool (MMAT) version 2018 for information professionals and researchers. Education for Information, 2018. 34(4): p. 285–291.

17. Puttick, R.L., J., Standards of evidence: an approach that balances the need for evidence with innovation. 2013.

18. (UTRIP), I.f.R.a.D.U., The role of online and online peer support for young people who self-harm. 2012.

19. Brow, L.K., Online self-injury message boards: The role of social support for young adults. Dissertation Abstracts International: Section B: The Sciences and Engineering, 2016. **76**(10-B(E)): p. No-Specified.

20. Lavis, A. and R. Winter, #Online harms or benefits? An ethnographic analysis of the positives and negatives of peer-support around self-harm on social media. Journal of Child Psychology and Psychiatry, 2020. 61(8): p. 842–854.

21. Smithson, J., et al., Membership and boundary maintenance on an online self-harm forum. Qualitative health research, 2011. 21(11): p. 1567–75.

22. Sharkey, S., et al., Supportive interchanges and face-work as ‘protective talk’ in an online self-harm support forum. Commun Med, 2012. 9(1): p. 71–82.

23. Corcoran, J., A. Mewse, and G. Babiker, The Role of Women’s Self-injury Support-Groups: A Grounded Theory. Journal of Community & Applied Social Psychology, 2007. 17(1): p. 35–52.

24. Murray, C.D. and J. Fox, Do Internet self-harm discussion groups alleviate or exacerbate self-harming behaviour? AeJAMH (Australian e-Journal for the Advancement of Mental Health), 2006. 5(3): p. 1–9.

25. Haberstroh, S. and M. Moyer, Exploring an online self-injury support group: Perspectives from group members. Journal for Specialists in Group Work, 2012. 37(2): p. 113–132.

26. Boyce, M., C. Munn-Giddings, and J. Secker, “’It is a safe space’: Self-harm self-help groups”. Mental Health Review Journal, 2018. 23(1): p. 54–63.

27. Whitlock, J.L., J.L. Powers, and J. Eckenrode, The virtual cutting edge: the internet and adolescent self-injury. Developmental psychology, 2006. 42(3): p. 407–17.

28. Sharkey, S., et al., Supportive interchanges and face-work as ‘protective talk’ in an online self-harm support forum. Communication & medicine, 2012. 9(1): p. 71–82.

29. Frost, M., L. Casey, and N. Rando, Self-Injury, Help-Seeking, and the Internet: Informing Online Service Provision for Young People. Crisis, 2016. 37(1): p. 68–76.

30. Aguirre Velasco, A., et al., What are the barriers, facilitators and interventions targeting help-seeking behaviours for common mental health problems in adolescents? A systematic review. BMC psychiatry, 2020. 20(1): p. 293.

31. Kitchener, B., Jorm, AF, Mental health first aid training for the public: evaluation of effects on knowledge, attitudes and helping behavior. BMC Psychiatry, 2002. 2(10).

32. Aref-Adib, G., McCloud, T., Ross, J., O’Hanlon, P., Appleton, V., Rowe, S., Murray, E., Johnson, S., Lobban, F., Factors affecting implementation of digital health interventions for people with psychosis or bipolar disorder, and their family and friends: a systematic review. The Lancet Psychiatry, 2019. 6(3): p. 257–266.

